# Far from the gut but still relevant? Circulating bacterial signature is linked to metabolic disease and shifts with metabolic alleviation after bariatric surgery

**DOI:** 10.1101/2021.03.25.21254332

**Authors:** Rima M. Chakaroun, Lucas Massier, Anna Heintz-Buschart, Nedal Said, Joerg Fallmann, Alyce Crane, Tatjana Schütz, Arne Dietrich, Matthias Blüher, Michael Stumvoll, Niculina Musat, Peter Kovacs

## Abstract

**Background:** The microbiome has emerged as an environmental factor contributing to obesity and type 2 diabetes (T2D). While the majority of studies have focused on associations between the gut microbiome and metabolic disease, increasing evidence suggests links between circulating bacterial components (i.e. bacterial DNA) and cardiometabolic disease as well as blunted response to metabolic interventions such as bariatric surgery. In this aspect, thorough next generation sequencing based and contaminant aware approaches are lacking. To address these points, we tested whether bacterial DNA could be amplified in the blood of subjects with obesity and high metabolic risk under strict experimental and analytical control to minimize bacterial contamination. Moreover we characterized a bacterial signature associated with the individual metabolic risk and explored its dynamics alongside metabolic improvement after bariatric surgery.

**Methods:** Subjects undergoing elective bariatric surgery were recruited into sex and BMI matched subgroups with (n=24) or without T2D (n=24). Bacterial DNA in the blood was quantified and prokaryotic 16S rRNA gene amplicons were sequenced. A contaminant aware approach was applied to derive a compositional microbial signature from bacterial sequences in subjects with and without T2D and within subjects at baseline and at three and twelve months post bariatric surgery. We modelled associations between bacterial load and composition with host metabolic and anthropometric markers. We further tested whether compositional shifts were related to weight loss response and T2D remission after bariatric surgery. Lastly, Catalyzed Reporter Deposition (CARD) - Fluorescence *In Situ* Hybridization (FISH) was employed to visualize bacteria in blood samples.

**Results:** Contaminant aware classification of bacterial 16S rRNA sequences allowed the derivation of a blood bacterial signature, which was associated with metabolic health. Based on bacterial phyla and genera detected in the blood samples, a metabolic syndrome classification index score was derived and shown to robustly classify subjects along their actual clinical group. T2D was characterized by decreased bacterial richness and a loss of genera associated with improved metabolic health. Moreover, circulating bacterial load was significantly associated with metabolic health and increased after bariatric surgery. Weight loss and metabolic improvement following bariatric surgery were associated with an early and stable increase of these genera in parallel with improvements in key cardiometabolic risk parameters. CARD-FISH allowed the detection of living Bacteria in blood samples in obesity.

**Conclusions:** We show that the circulating bacterial signature reflects metabolic disease and its improvement after bariatric surgery. Our work provides contaminant aware evidence for the presence of living bacteria in the blood and suggests a putative crosstalk between components of the blood and metabolism in metabolic health regulation.

## Introduction

The major health burden of obesity is largely attributable to its associated diseases such as type 2 diabetes (T2D), the metabolic syndrome (MetS) and cardiovascular diseases. Along know risk factors, such as genetic predisposition, poor diet and lower physical activity, shifts in the gut microbial composition have been shown to contribute to metabolic inflammation at the advent of obesity and T2D [1–5]. Beyond composition, low gut bacterial diversity has been associated with increased obesity, insulin resistance, dyslipidemia and increased inflammation [6, 7]. More recently, low gut bacterial load has been identified as a key driver related to chronic inflammatory states such as Crohn’s disease [8]. Converging evidence further points to an important role of the gut microbiome as a therapeutic target and prognostic marker: Weight loss interventions, such as diet and bariatric surgery, profoundly affect the gut microbiota composition, leading to an increased bacterial gene count and bacterial richness. This, in turn, is associated with a decrease in inflammatory markers and an increase in insulin sensitivity up to complete remission of T2D [9–11]. Accordingly, weight loss interventions are less effective in improving insulin resistance and inflammatory state in patients with low bacterial gene diversity [7].

Although most evidence has been submitted for the gut microbiome, host tissues – including blood [12], liver and adipose tissue [13, 14]– have been shown to accommodate microbial consortia finally accessible through culture independent techniques. Few studies further linked increased bacterial DNA load in the circulation with the incidence of T2D [15, 16] and cardiovascular disease [17]. Moreover, bacterial signature in the blood has been linked to the circulatory compartment it derives from (i.e. systemic vs. portal circulation) [18], systemic inflammation [18], T2D presence [13, 14, 19] and metabolic disease severity [20]. Similarly, there is mounting evidence for diagnostic application of blood based bacterial DNA highlighted in a recent work employing contaminant-aware approaches to show that it can discriminate between multiple types of cancer and is highly dependent on disease severity [21]. On the other hand, only one study contemplated the interplay between metabolic interventions and circulating bacterial markers: Subjects with an established bacterial DNA translocation based on qPCR detection did not experience a remission of T2D or significant improvement in insulin sensitivity despite significant weight loss after bariatric surgery [22]. These results advocate a closer look at blood-borne bacterial DNA and bacterial signatures and their role in health and disease.

It is worth noting that studies on bacterial load and composition in low bacterial biomass environments such as the blood are subject to technical biases including highly underestimated environmental contamination [23]. To our knowledge, only few studies have actively controlled for contamination so far [13, 14, 21], and only two included a bioinformatic contaminant aware approach [13, 14]. We therefore applied a contamination aware approach to test the hypothesis that the presence, composition and load of bacterial DNA in the blood reflects obesity, metabolic risk factors and inflammatory burden as well as weight loss associated changes in anthropometric and metabolic parameters after bariatric surgery.

## Research design and methods

### Characterization of study participants and sample collection

64 subjects were recruited at the University of Leipzig Medical Center, Germany, after matching for BMI and sex differing on T2D status with 32 subjects having no T2D and 32 with T2D according to ADA-criteria (19). However, 16 subjects had to be excluded from longitudinal comparisons due to missing follow-ups in at least one time point (n=15) or missing follow-up phenotypes (n=1). If not indicated otherwise, 48 subjects (24 with T2D, 24 without) were included in all analyses (**table 1**- baseline cohort characteristics of subjects with complete follow-up (n=48), **supplementary table 1**- baseline cohort characteristics with initial matching (n=64)). Individuals fulfilled the following inclusion criteria: 1) Eligibility to bariatric surgery according to international clinical guidelines (BMI >= 40 kg/m^2^, or >= 35 kg/m^2^ with at least one obesity associated metabolic disease) and internal clinical multidisciplinary panel, 2) no chronic or acute inflammatory disease as determined by blood cell counts and CRP or clinical signs of infection, 3) no evidence of coronary or peripheral artery disease, 4) no known thyroid dysfunction, 5) no antibiotics intake three months prior to the study visit, 6) no pregnancy or nursing, and 7) no NSAIDs intake within 78 h prior to the study visit.

**Table 1.**
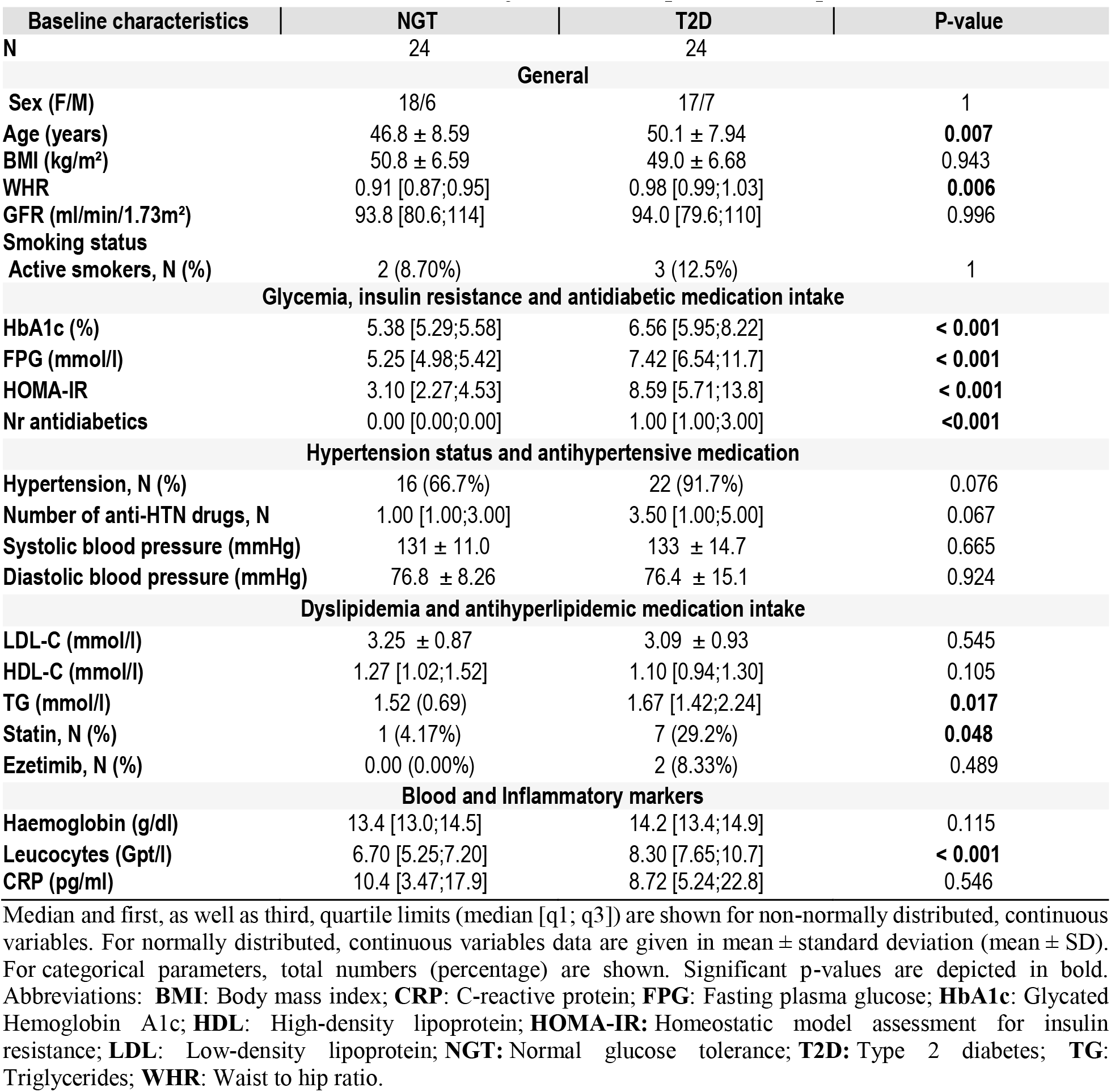
Baseline cohort characteristics of subjects with complete follow-up

Subjects were extensively phenotyped before bariatric surgery and at month three and twelve post bariatric surgery, including clinical and anthropometric assessment in conjunction with evaluation of patient medical history and physical examinations. Among other parameters, blood pressure, waist and hip circumference measurements as well as bioimpedance measurement for body composition analysis, including automatic calculation of visceral fat rating, (BC-418 MA, Tanita, Tokyo, Japan) were assessed. Obesity associated diseases such as T2D [24], hypertension [25] and hyperlipidemia [26] were defined according to medical association definition of the specific disease. Metabolic syndrome (MetS) was defined according to international conventions [27]. Excess BMI loss (EBL) was calculated as follows: EBL = (BMI baseline - BMI 12 months) / (BMIbaseline- 25) * 100 [28] and poor response was defined as an EBL <50%, whereas a good response was defined as an EBL >60% (adapted from [28]) (**supplementary table 2**– phenotypes of good vs poor responders).

### Replication Cohort

To validate results from bacterial quantification and associations thereof with host parameters, data from a subgroup consisting of 62 subjects belonging to a previously described cohort from the same center were analysed (**supplementary table 3 – phenotypes replication cohort**). Phenotyping procedures and cohort description of the whole cohort are available elsewhere [14].

### Samples preparation

Blood samples were collected after an overnight fast at each time point, and EDTA-blood samples were collected for DNA isolation and stored at −20°C. Fasting plasma glucose was measured using the hexokinase method, HDL- and LDL-cholesterol were measured using enzymatic assays and insulin was measured using chemiluminescence assay according to standard laboratory procedures. C-reactive protein (CRP) was measured using an Image Automatic Immunoassay System (Beckman Coulter). All measurements were performed according to routine laboratory procedures. Homeostasis Model Assessment of Insulin Resistance (HOMA-IR) was calculated as previously described [29]. Lipopolysaccharide binding protein (LBP) was measured using the HK315 HUMAN LBP ELISA Kit (Hycult Biotech, Uden, Netherlands) according to the manufacturer’s recommendations. Samples and follow up availability dictated the numbers of used samples for analyses at each step as illustrated in **supplementary figure 1**.

### Bacterial DNA extraction, quantification and amplification for sequencing

#### Bacterial DNA extraction

To minimize contamination, all steps were performed under a sterile class II laminar flow hood using aseptic measures including the use of one-way lab coats, elbow-length gloves as well as facemasks. All non-organic liquids and required materials were subjected to at least 120 minutes of UV-radiation. Furthermore, negative controls consisting of UV-treated PBS (filled in EDTA vials using venipuncture ware from the same batch to simulate venous puncture) were included and carried through all experimental procedures, including sequencing.

DNA was extracted from 200 µl whole EDTA-blood using the QIAMP Blood MiniKit (Qiagen, Germany) following the manufacturer’s recommendations with an additional lysozyme step at 37 °C overnight (0.25 mg/ml, L7615, Sigma-Aldrich, MO, USA). DNA yield, integrity and quality were assessed using Quant-iT PicoGreen dsDNA kit (Invitrogen, CA, USA) and Nanodrop 2000 spectrometer (Thermo Fisher Scientific, MA, USA).

#### Quantification of bacterial 16S rRNA gene

Bacterial DNA was quantified by qPCR amplification of the bacterial 16s rRNA gene using previously published primers (F_Bact 1369: 5’-CGGTGAATACGTTCCCGG-3’ and R_Prok 1492:5’- TACGGCTACCTTGTTACGACTT-3’) [17, 30, 31]. All qPCR reactions were performed in duplicate each using 50 ng of whole extracted DNA, prepared in a PCR clean room and run on a LightCycler 480 (Hoffmann-La Roche, Basel, Switzerland) with the following conditions: 50 ° C for 2 min, 95 °C for 10 min and 40 cycles of denaturation at 95 °C for 15 s, annealing at 60 °C for 30 sand extension at 72 °C for 30 s. The amount of amplified bacterial DNA was calculated using mean Cp values for each duplicate against a standard curve from E. coli JM 109 DNA dilutions (Promega, MA, USA), which included seven duplicates ranging from 1.25 fg to 0.2 ng total bacterial DNA. Quantification of the analyzed samples was performed in three runs with R^2^ ≥0.995 and a mean PCR efficiency of 2.06 ± 0.03. Analysis was conducted according to the Livak method [32]. Results were in concordance with a commercially available kit (Zymoresearch, CA, USA) (n=13, r=0.74, p=0.004) and proved more sensitive (ΔCt=3.9). Obtained concentrations were normalized to the total concentration of extracted DNA as well as the used blood volume and is given as pg bacterial DNA per µg of isolated DNA. This is due to the variation of amounts of total extracted DNA from the same 200 µl blood volume for each sample, which ranged from 12.7 - 98.2 ng/µl. Blood bacterial quantity in the replication cohort (**supplementary table 3**) was independently measured according to the same protocol. To overcome inter-assay variability, blood bacterial load was standardized (z-score transformation) within the cohorts and analyses were conducted on standardized bacterial quantities for each cohort separately as well as both of them combined.

#### 16S rRNA gene sequencing analysis

After extensive testing of combinations of primers and polymerases for amplification of prokaryotic / bacterial 16S rRNA variable regions [14], V4-V5 primers adapted from the Ribosomal Database Project (RDP) (V4-F: 5’-ACTGGGCGTAAAGCG-3’; V5-R: 5’-CCGTCAATTCCTTTGAGTTT-3’) were used [33]. PCR reactions were prepared in triplicate in a sterile laminar flow hood and performed using 50 ng total DNA in a total reaction volume of 25 µl containing 1.25 µl of each primer (10µM), 12.5 µl 2x Q5 Reaction Buffer (including Q5^®^ High-Fidelity DNA Polymerase, New England BioLabs, MA, USA) and 6.25 µl UV-ed PCR-grade water. The reaction was carried out on a LightCycler 480 (Hoffmann-La Roche, Basel, Switzerland) under the following conditions: 98 °C for 2 min and 40 cycles of denaturation at 98 °C for 30 s, annealing at 58 °C for 30 s and extension at 72 °C for 30 sec. Each PCR reaction included at least 3 negative controls in addition to the extraction controls (blank controls) and the absence of detectable PCR products in these negative controls was confirmed by gel electrophoresis. Regardless of this, negative controls for each run were pooled and carried through sequencing to be used for contaminant identification. Replicate amplicons were pooled and purified using Agencourt Ampure magnetic purification beads (Beckman Coulter Indianapolis, IN, USA) according to the manufacturer’s protocol to remove short amplification products and primer dimers. DNA amounts were quantified using Quant-iT PicoGreen dsDNA kit (Invitrogen, CA, USA). Library preparation and paired-end sequencing were performed commercially (BGI Genomic, Shenzhen, China) on Illumina Hiseq2500 technology and using custom fusion-primers in a one-step PCR approach.

### Bioinformatics and statistical analyses

#### Statistical analyses

Statistical analyses were performed in R v3.5.0 (R Development Core Team, 2008). Prior to statistical testing, distribution as well as single test assumptions were checked and statistical tests were chosen accordingly. For cross-sectional comparisons between two groups, Wilcoxon signed-rank-test was used for non-normally distributed variables according to Shapiro-Wilk testing, whereas unpaired Student’s t-test was used to compare normally distributed variables. Median and first as well as third quartile limits are depicted for non-normally distributed variables, whereas for normally distributed continuous variables, data are given in mean ± standard deviation (SD). Friedman-test was used to compare dependent groups at different time points. Categorical parameters were analyzed using the Chi-square test. Bivariate correlation analyses were performed using spearman’s rank correlation test. Figures were generated using ggplot2 [34] v3.1.0, ggpubr v0.2 [35], and corrplot v.0.84 [36] as well as phyloseq v1.26.1 [37] and relabeled in Adobe Illustrator (Adobe Inc., CA, USA). A p-value threshold of 0.05 was used to depict statistical significance. Analyses were adjusted for multiple hypotheses *ad modum* Benjamini-Hochberg, in which case a False-discovery-rate (FDR) of *Padj* <0.05 was considered significant.

#### Processing of 16S rRNA reads and amplicon sequence variants (ASV) clustering

A total of 12,857,649 Illumina Hiseq quality filtered paired end reads were obtained from BGI, with samples having a median read count of 63,055 [9661 – 64,630]. Preprocessing and quality filtering consisted of removing of reads with a certain proportion of low quality (20) bases (20% of read original length), contaminated by adapter (5 bases overlapped by reads and adapter with maximal 3 bases mismatch allowed), with ambiguous bases and with low complexity (reads with 10 consecutive same base). Subsequently, quality was checked using Multiqc [38] and data was imported to QIIME2 [39] V2019.1 for **analysis (**supplementary file 1 for Qiime2 commands – DOI reserved and citation to follow **acceptance**). Denoising, dereplication, merging and chimera-filtering as well as ASV inference were done in one step using the DADA2 [40] plugin in QIIME 2 resulting in 40269 ± 6033 non-chimeric reads with an average read length of 329 bp. Reads were truncated at a length of 242 bp. For phylogenetic analyses, primer-truncated and quality trimmed reads were used to create an alignment using mafft [41]. The alignment was masked to remove uninformative highly variable regions and a rooted tree was generated using the align-to-tree-mafft-fasttree plugin. For taxonomic classification a scikit-learn [42] naive Bayes classifier was created against the taxonomic classification from ARB-SILVA [43] 132 release (99% OTU data set), which was trained for the used primers. Only reads mapped to bacterial taxonomy were retained. This resulted in a total of 2860 bacterial ASVs with a total frequency of 7,784,509. Derived taxonomy, tree and feature table were imported into phyloseq [37] v1.26.1 and all subsequent analyses were performed in R v 3.5.0 0 (R Development Core Team, 2008).

#### Bacterial contaminants identification in blood samples

A bioinformatic decontamination step using the Decontam [44] package v1.2.1, which is intended for the identification of contaminants in low bacterial biomass samples, was added. To identify contaminant ASVs, the “prevalence” method was applied. A binomial distribution with low scores for low prevalence ASVs was found around 0.1 and high prevalence ASVs with higher confidence increasing around 0.175, which led us to select a data driven contaminant score of 0.175 instead of the default classification score 0.1 (**supplementary figure 2**). Using these parameters, 114 features were classified as potential contaminants, and removed, leading to a total of 2746 ASVs (**supplementary file 2**: contaminating ASVs at Decontam Score 0.175, **supplementary file 3**: taxonomy in samples and negative controls, **supplementary file 4:** taxonomy in samples after removing contaminant sequences at a contaminant score of 0.175). The pruned phyloseq object was then used for subsequent analyses. We moreover added a more stringent decontamination step using a contaminant score of 0.5 leading to flagging of each of the 172 ASVs appearing in the negative samples as contaminants. While ASVs derived from both decontamination scores led to similar results in associations of taxonomy with host variables (data not shown), the data driven decontamination at a lower score was more likely to influence differential abundance analyses. This led us to use a more stringently decontaminated taxonomy while performing differential abundance testing to avoid spurious observations. The stringently decontaminated taxonomy included a total of 2688 ASVs. (**supplementary file 5:** contaminating ASVs at Decontam Score 0.5, **supplementary file 6** : taxonomy in samples after removing contaminant sequences at a contaminant score of 0.5).

#### Analyses of bacterial composition and its association with host phenotypes

Alpha diversity measures (Shannon diversity, observed richness) were calculated and RDA on Bray-Curtis-dissimilarity distances was conducted to identify host covariates contributing to bacterial community composition in the blood in vegan [45] v2.5-4. Samples with missing observations (NAs) in the covariates were eliminated from the dataset prior to analyses. Variables used in the RDA model included: T2D Status, Sex, Metformin use, PPI use, timepoint, MetS status, diabetes alleviation after surgery, EBL, surgical procedure, BMI, systolic blood pressure, eGFR, CRP, HbA1c, HDL-cholesterol, LDL-cholesterol, triglycerides, total fat mass in %, visceral fat ratio, number of antidiabetics, number of antihyperlipidemic drugs, number of antihypertensive drugs, white blood cells count, age, bacterial load and waist-to-hip-ratio. We further used automatic stepwise modelling and model selection using the ordistep approach both ways with 999 permutations to identify the most significant covariates contributing to community composition. Microbiome [46] v1.4.2.was used to perform correlation analyses between relative taxa abundances and host variables of interest applying Spearman’s rank correlation.

A random forest classification (randomForest [47] v4.6-14) approach on both phylum and genus level abundances was used to develop classification indices for MetS for subjects at baseline. For phylum and genus level NGS derived read counts from MetS and non-MetS subjects where normalized by the total read count of the corresponding phylum or genus and used as features for training of a random forest classifier. The classification index predicted by the machine ranges between 0 and 1 and corresponds to the out-of-bag predicted probability of being classified as belonging to the class of interest (i.e. having MetS) with higher scores indicating higher probability for having MetS. Performance of classification indices were quantified using receiver operating characteristic curve and AUC using pROC [48] v1.14.0. Cross validation was performed using the rfUtilities package [49, 50] v2.1-5. Specifics of random forest classification are as follows: Number of trees 500; Number of variables tried at each split 11; OOB estimate of error rate at genus level 21.3%, at phylum level 29.8%; Classification accuracy for model at genus level: users accuracy 100, producers accuracy: 79.1, Model kappa=0.147, Model OOB Error=0.204, Model Error variance= 4.7 x1 0^-5^; Classification accuracy for model at phylum level : users accuracy 91.2, producers accuracy: 79.5 Model kappa=0.1357, Model OOB error 0.25, Model error variance = 7.7 x 10^-4^).

Differential abundance analyses for preassigned dichotomous groups were performed using DESeq2 [51] v1.22.2. Size factors for count data were calculated using the poscounts estimator. Dispersions estimates were performed using the DESeq command, a local regression model was used to fit and test the data and the negative binomial Wald Test was used to test differential abundance (log2 fold change) and significance. Results are reported for differentially abundant taxa with a significance *Padj* < 0.01 after multiple hypotheses correction *ad modum* Benjamini-Hochberg. Specifically, for the comparison between T2D remission and no remission following bariatric surgery, only subjects with T2D at baseline were retained in the analyses (**supplementary figure 3**).

### Catalyzed Reporter Deposition (CARD) - Fluorescence In Situ Hybridization (FISH)

#### Blood sample collection and cell separation

To visualize and quantify bacteria in the blood, samples from one patient post bariatric surgery ( male, 38 years, currently 60 months post bariatric surgery, current BMI 30 kg/m^2^, no T2D, no MetS, initial BMI 61.9 kg/m^2^, EBL 54.5%) and a healthy control (male, 50 years, BMI, no known diseases, no medication).were collected before and 30 min after mixed meal intake on EDTA 1.6 mg/mL to avoid coagulation and kept overnight at 4°C in the fridge. Density gradient centrifugation was performed in an initial step to separate microbial cells from blood cells. Thus, 5 mL of 80% Nycodenz solution were added at the bottom of a tube containing 5 mL of blood, followed by centrifugation at 13000 rpm, 4°C for 2 h. After density gradient centrifugation, individual layers consisting of supernatant and a Nycodenz layer could be separated. Each layer was individually collected and subjected to serial filtration through two consecutive 3µm and one 0.22µm pore size polycarbonate filters (GTTP type, 0.2µm pore size PC membrane, 25mm diameter, Merck Millipore, Germany). After filtration, all filters were washed in 1× UV-ed PBS buffer and were immersed in 4% paraformaldehyde solution (PFA) in 1×UV-ed PBS buffer for 2h at room temperature. Following chemical fixation, filters were washed in 1× UV-ed PBS buffer, dehydrated in 80% ethanol, air dried and stored at 4°C for CARD-FISH procedure. **CARD-FISH** was performed following the standard protocol [52] with slight modifications. Permeabilization in Lysozyme solution (Sigma-Aldrich, MS, USA) (10 µg mL-1) in 0.1M Tris-HCL (pH=7.8) and 0.05M EDTA (pH=8) buffer was done for 30 min at 37°C followed by 0.01M HCL for 10 min at room temperature (RT) with washing steps of 1 min at RT in ultrapure water in between treatments. Hybridization of filter pieces belonging to both 3µm and 0.2µm filters took place for 3 h at 46°C in a pre-warmed hybridization buffer containing 0.9M NaCl, 20mM Tris–HCl (pH=7.5), 10% (w/v) dextran sulfate,0.02% (w/v) SDS, 35% (v/v) formamide (Fluka, Waltham, USA) and 1% (w/v) blocking reagent (Boehringer, Mannheim, Germany). The HRP-labelled probe applied is specific for Bacteria (EUB 338, Amann et al., 1990) and was used at a concentration of 0.166 ng mL^−1^ (HRP-probe stock solution of 50 ng mL^−1^ diluted 1:300 v/v in hybridization buffer). Following hybridization, filter pieces were incubated in 50 mL of pre-warmed washing buffer containing 70mM NaCl, 5mM EDTA (pH=8.0), 20mM Tris–HCl (pH=7.5), and 0.01% SDS for 15 min at 48°C. After washing, filters were incubated for 15 min at RT in 1× PBS (pH=7.6) to equilibrate the HRP-labeled probe. Subsequently, tyramide deposition was performed by incubating the filters for 20 min at 46°C in the dark in amplification buffer containing 1× PBS, 2M NaCl, 0.1% (w/v) blocking reagent, 10% (w/v) dextran sulfate, 0.0015% (v/v) H_2_O_2_, and 1 µg mL^−1^ Alexa 594- labeled tyramides (ThermoFisherScientific, MA,USA). Afterwards, the hybridized filters were rinsed in 1× PBS for 10 min at RT followed by staining with 4’,6-diamidino-2-phenylindole (DAPI) 1µg mL-1 for 10 min at RT, washing in ultrapure water, air dried and embedded in mounting medium (a mixture of low fluorescence glycerol mountant (Citifluor AF1, London) and mounting fluid Vecta Shield (Vecta Laboratories, CA, USA) in a 4:1 v/v ratio and stored at −20°C prior to imaging.

#### Microscopic Investigation

Approximately 20-30 randomly selected fields of view (each of 15130.8 µm^2^) were imaged on each hybridized filter piece from 3µm and 0.2µm pore size hybridized filters. The microscopic evaluation of the hybridized filters was done using a fluorescence microscope (Imager. Z2, Zeiss, Germany) with 20X air (numerical aperture (NA= 0.5) and 63X oil (NA= 1.4) objectives. On the 3µm filters we observed predominantly un-hybridized blood cells of different sizes for both Supernatant and Nycodenz filtrated samples while hybridized bacterial cells were constantly found on the 0.22µm filters of the Nycodenz filtrated samples, occasionally in very low abundances also on the 0.22 filters of the Supernatant filtrated samples (data not shown). Bacterial counts were performed only on the 0.22µm filters from the Nycodenz filtrate layer on 20 to 22 randomly selected fields of view..

## Results

### Contaminant aware analyses allow the derivation of a core blood bacterial signature

After filtering and removing contaminating ASVs (**supplementary file 2**), a distinct bacterial profile remained consisting of 20 phyla (**supplementary file 4**). Of the 2746 detected ASVs, 65 could not be assigned at the phylum level (2.4%). Assigned taxonomy was dominated by *Proteobacteria* (59.0%), *Firmicutes* (12.4%), *Patescibacteria* (11.97 %), *Cyanobacteria* (8.9%), and *Actinobacteria* (2.3%), whereas 5.41% of ASVs belonged to various other phyla.

21.6% of all ASVs (592) could not be assigned at the genus level. Assigned ASVs belonged to 314 genera and the most abundant genera included *Aliterella, Anoxybacillus, Lactobacillus* and *Sphingomonas* from the three dominant phyla (**fig 1A**). The core bacterial signature consisting of 10 phyla and 120 genera with an overall abundance of more than 10% in the dataset could be recovered at each timepoint, although quantitative changes could be tracked over time and within disease and response groups (see further results) (**fig 1B,C**).

**Figure 1.**
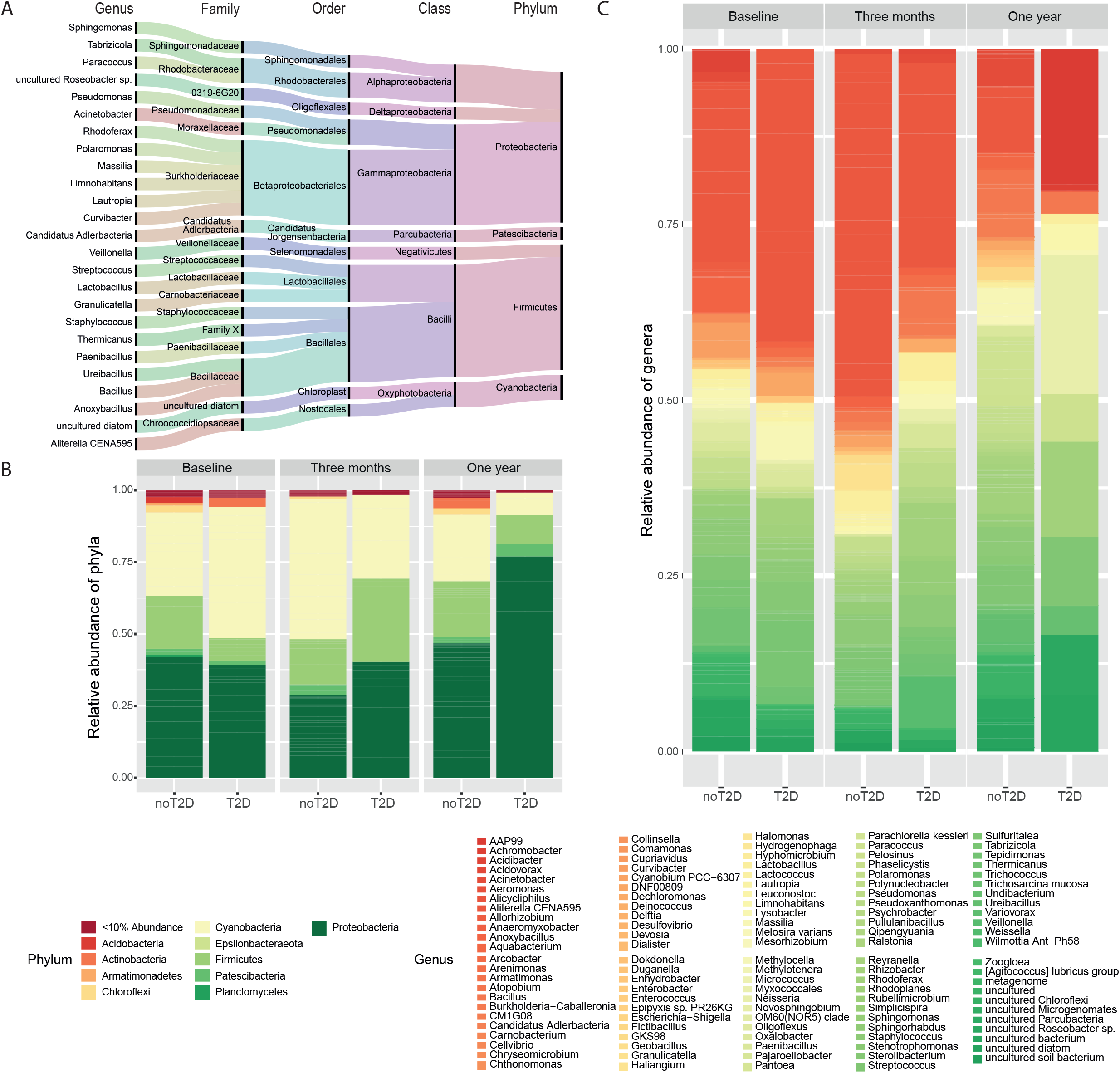
contaminant aware blood bacterial signatures at a decontam score of 0.175 **(A)** Top 25 genera and their respective taxonomic classification. The bandwidth is proportional to the amount of ASVs within each Phylum, genera are ordered from bottom to top within each family according to alphabetical order, taxonomy was generated with all available baseline samples (n=64) and follow-up samples at three months (n=48) and one year (n=48) **(B)** Relative abundance of phyla within samples with follow-up sequencing at all timepoints: Baseline (n=48), three months (n=48) and one year (n=48) after bariatric surgery. Phyla with an abundance within the whole dataset of less than 10% are flagged under the category ‘<10% Abundance’. Taxonomy is shown for matched subjects with and without T2D (n for each group =24) at each timepoint separately**. (C)** Relative abundance of core genera belonging to the phyla with an abundance of >10% within samples with follow-up sequencing at all timepoints: Baseline (n=48), three months (n=48) and one year (n=48) after bariatric surgery. Taxonomy is shown for matched subjects with and without T2D (n for each group =24) at each timepoint separately.

### Bacterial community composition is influenced by medication and reflects metabolic health

Twenty-seven available host variables (**supplementary tables 4 and 5**) were fit onto genus level RDA in all available samples with no missing data (n=101). RDA showed that 39.7% of the observed variance could be explained by the collected host variables (R^2^= 0.3969, R^2^adj=0.162, p-values of Anova = 0.004). Associations with RDA ordination on Hellringer transformed abundances were most notable for timepoint (envfit; R^2^= 0.2184, p=0.001), white blood cell count (envfit: R^2^ = 0.2531, p=0.001), BMI (envfit; R^2^= 0.2219, p=0.001), total fat mass (envfit; R^2^= 0.2090, p=0.001), bacterial quantity (envfit; R^2^= 0.1788, p=0.001) and triglycerides (envfit; R^2^= 0.1560, p=0.001). Several other significant associations with bacterial composition were observed for general host characteristics such as age, T2D status, MetS status, sex, anthropometric variables such as waist-to-hip-ratio and visceral fat rating and markers of metabolic control including HDL-cholesterol and HbA1c as well as inflammation markers like CRP and bacterial DNA burden (**supplementary table 4**: envfit output for 27 variables on genera level RDA, **supplementary figure 4**). Similar results were observed at ASV levels as well (**supplementary table 5**).

Interestingly, while the number of antihypertensive, antidiabetic and antihyperlipidemic medication was not correlated with ordination, the use of metformin and PPI significantly contributed to variance in composition (Metformin R^2^= 0.0380, p=0.022, PPI R^2^=0.0674, p=0.001, **supplementary table 4 and 5**).

### Blood bacterial signature allows robust classification of subjects with metabolic syndrome

Cardiovascular disease and mortality are increased in subjects with MetS [53] even in the absence of T2D. Because MetS status was significantly associated with bacterial composition beyond single risk factors used in the MetS definition [27] (**supplementary tables 4 and 5**), we sought to investigate if bacterial composition could predict MetS status. For this, we applied random forest classification on the genus as well as phylum level abundances (20 phyla and 314 genera) to develop a classification index. The resulting MetS classification index (MetS-I_genus_ and MetS-I_phylum_) could robustly classify subjects along their actual clinical group at baseline and performed better at genus than at phylum level (AUCgenus=0.816, 95% CI: 0.661-0.9705 (DeLong) and AUCphylum=0.740 95% CI: 0.589-0.89 (DeLong) (**fig 2A-D**). MetS-I at both genus and phylum levels at baseline was correlated with several markers related to obesity, visceral fat distribution, insulin resistance and inflammation from all timepoints, but was negatively associated with bacterial DNA load in the blood (**fig 2E, F**). Largest associations with host variables were seen for MetS-Igenus, which was positively correlated with anthropometric markers related to obesity and insulin resistance such as total fat mass, waist circumference and WHR. It was also positively associated with inflammation reflected in white blood cells count, CRP and LBP as well as metabolic markers related to obesity and insulin resistance such as HOMA-IR, HbA1c, glucose, insulin and uric acid. The MetS-I_genus_ was further related to dyslipidemia and hypertension with positive correlations noted for triglyceride levels and number of antihypertensive drugs while being negatively associated with HDL-cholesterol. Moreover, both MetS-Igenus and MetS-Iphylum were negatively associated with bacterial quantity (**fig 2E,F**, **table 2)**.

**Figure 2.**
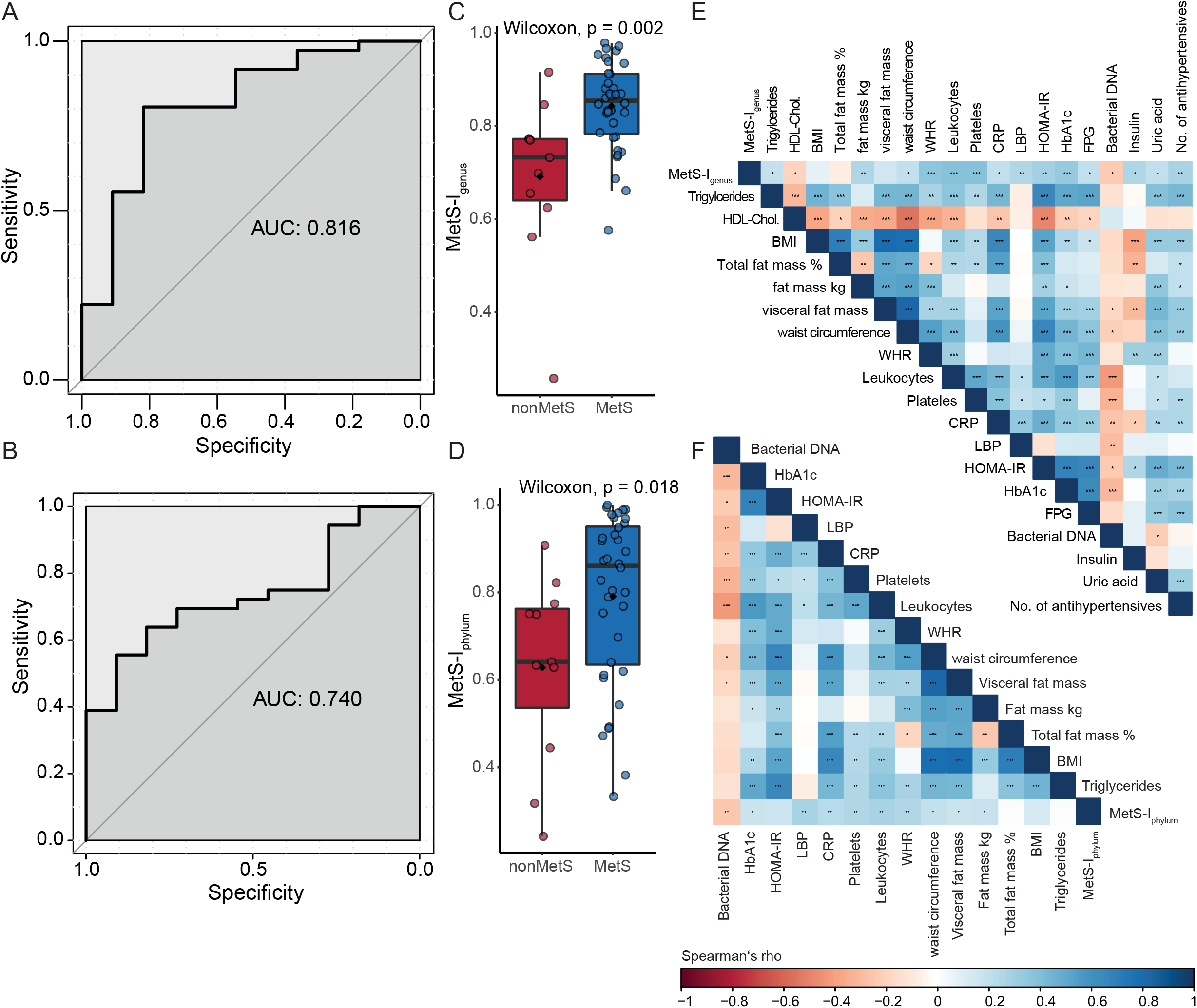
Bacterial blood signature derived metabolic syndrome classification index. Probability of being classified as subject with metabolic syndrome using random forest classification of Phyla and genera abundances (n Phyla=20, n genera=314, in 47 subjects – 36 with and 11 without MetS, missing n=1, nondiabetic, not unambiguously categorized) **(A, B)** corresponding Area under the curve for both genus and phylum level random forest models for metabolic syndrome prediction respectively **(C,D)** MetS-IGenus and MetS-IPhylum respectively in subjects with and without Metabolic syndrome, boxplots with Tukey- whiskers and mean (◆) are shown. Unpaired samples Wilcoxon signed-rank test is used to compare groups and nominal p-values are shown. **(E, F)** Spearman’s rank correlations of MetS-IGenus and MetS-IPhylum at baseline with host variables from all timepoints. Nominal significant values are indicated: *: p<0.05, **: p<0.01, ***: p<0.001.

**Table 2.**
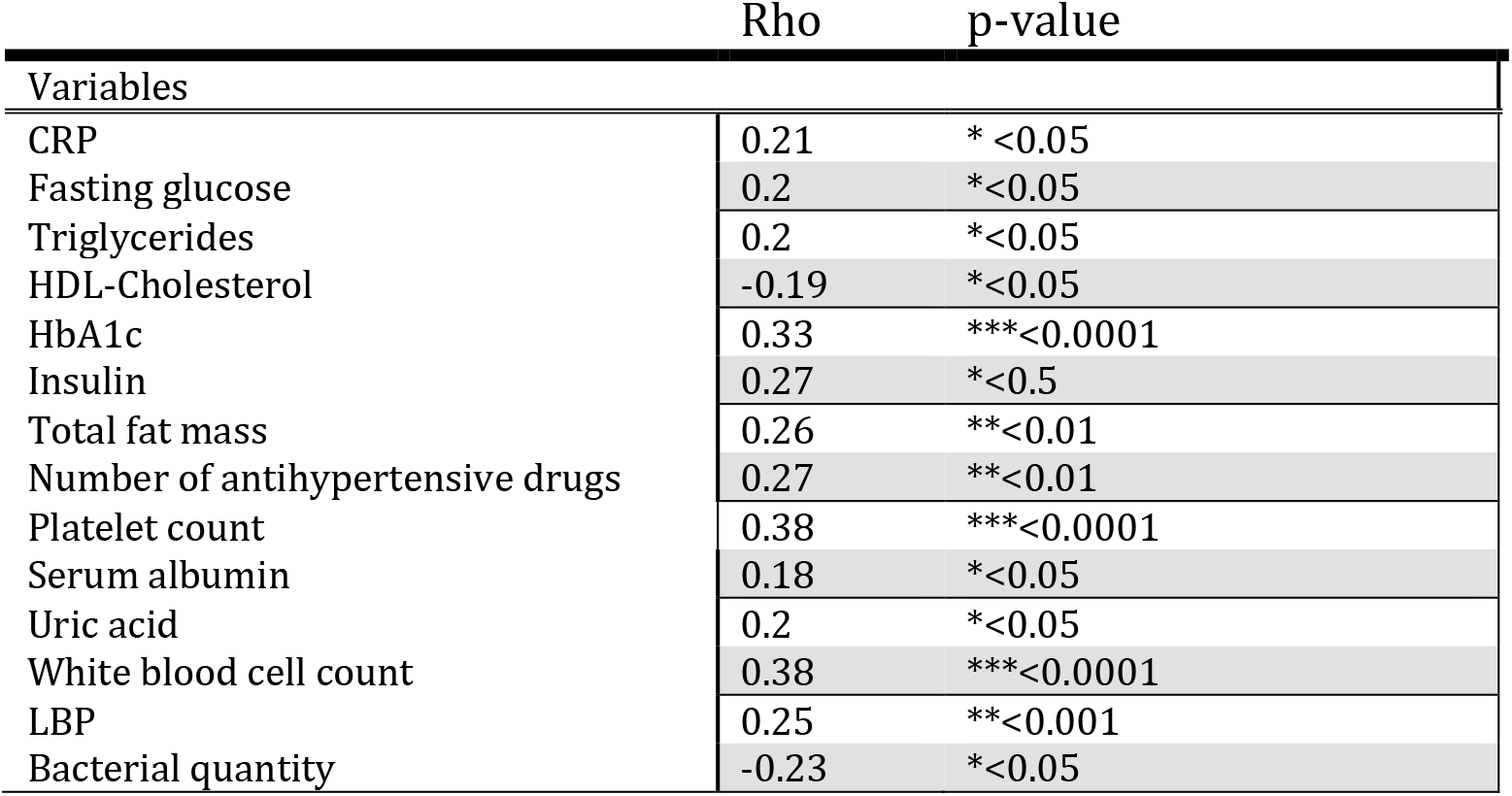
Spearman Correlations between MetS-Igenus at baseline with host variables (n=141: 47 subjects at 3 timepoints)

|  | Rho | p-value |
| --- | --- | --- |
| Variables |  |  |
| CRP | 0.21 | * <0.05 |
| Fasting glucose | 0.2 | * <0.05 |
| Triglycerides | 0.2 | * <0.05 |
| HDL-Cholesterol | -0.19 | * <0.05 |
| HbA1c | 0.33 | *** <0.0001 |
| Insulin | 0.27 | * <0.5 |
| Total fat mass | 0.26 | ** <0.01 |
| Number of antihypertensive drugs | 0.27 | ** <0.01 |
| Platelet count | 0.38 | *** <0.0001 |
| Serum albumin | 0.18 | * <0.05 |
| Uric acid | 0.2 | * <0.05 |
| White blood cell count | 0.38 | *** <0.0001 |
| LBP | 0.25 | ** <0.001 |
| Bacterial quantity | -0.23 | * <0.05 |

### T2D is characterized by a loss of health associated genera and a decreased ASV richness

We investigated bacterial alpha diversity according to T2D status: At baseline subjects with T2D (n=24) displayed significantly lower observed richness as compared to subjects without T2D (n=24) (mean observed richness = 28 ± 2.31 vs 19 ±2.10, p=0.039 in subjects without T2D and with T2D respectively; **fig 3A**). T2D was associated with significant shifts in several genera as compared to subjects without T2D: At baseline, significantly lower abundances of *Anoxybacillus*, *Duganella*, *Acidibacter* and *Chryseomicrobium* as well as *Sphingomonas* are found in T2D (**fig 3B**). Furthermore, timepoint seemed to be relevant for the differences in bacterial abundances between subjects with and with T2D (**supplementary figures 5A, B**). There were no significantly differentially abundant ASVs between T2D and nonT2D overlapping between baseline and one year, but the differences between subjects with and without T2D seemed to be shifting gradually. While subjects with T2D at baseline and three months showed a reduced abundance in *Sphingomonas*, a common feature seen between T2D at three months and one year is reduced *Pseudomonas* in T2D. Congruent T2D associated features seen at baseline and one year were reduced *Bacillaceae* and *Burkholderiaceae* (**supplementary file 7**). Some of the reduced genera in T2D were expectedly negatively associated with markers of metabolic disease and inflammation such as *Anoxybacillus* and *Sphingomonas* at baseline with leukocytes and blood pressure respectively (**fig 3C**) as well as *Paracoccus* with HOMA-IR and leukocytes and *Rhodoferax* with markers of obesity at one year post bariatric surgery (**supplementary figure 5C**). Other genera were surprisingly positively associated with BMI such as *Sphingomonas* (at months three and twelve) or with lipids and blood pressure such as *Acidibacter* (at baseline, months three and twelve) (**figure 3C, supplementary figure 5C**)

**Figure 3.**
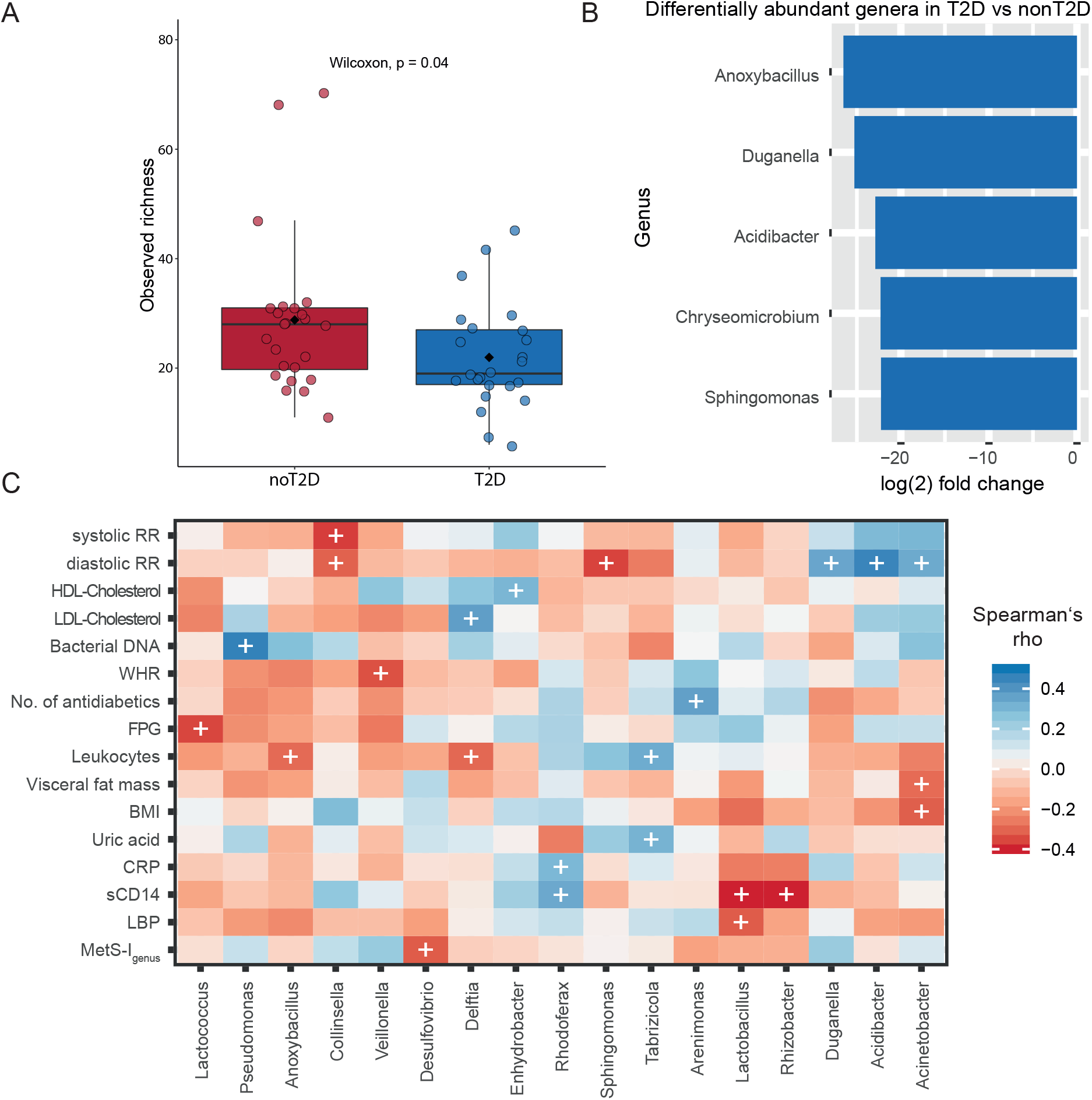
**(A)** Observed Richness between subjects with (n=24) and without T2D (n=24), boxplots with Tukey-whiskers and mean (◆) as well as median are shown. Unpaired samples Wilcoxon signed-rank test is used to compare groups and nominal p-value is shown. **(B)** Differentially abundant genera in subjects with T2D (n=24) compared to subjects without T2D (n=24) at baseline. Differential abundance is calculated at ASV level using taxonomy after stringent control for contamination at a decontam score of 0.5 and reported at genus level. **(C)** Spearman’s rank correlations of relative selected genera abundance with host parameters at baseline. Selection included genera seen to be differentially abundant between groups (i.e. T2D, good/poor responders and T2D alleviation vs no T2D alleviation). Only genera are shown with at least one significant correlation with host markers. (+) refers to p-value < 0.05. Color represents correlation strength (Rho) according to color legend. *Abbreviations*: HDL: high density lipoprotein; LDL: low density lipoprotein; WHR: Waist-hip-ratio; BMI: Body mass index; CRP: C-reactive protein; sCD14: soluble cluster of differentiation 14; LBP: Lipopolysaccharide binding protein; EBL: Excess BMI loss.

### Differential response in weight loss and glucose metabolism following bariatric surgery

As expected, bariatric surgery had a marked impact on BMI, body composition (WHR, total fat mass and visceral fat ratio), metabolism as well as inflammation as early as three months post procedure (**table 3**). From 24 subjects with T2D at baseline, nine subjects were still classified as T2D after three months, and only four after one year. From patients without T2D at one year, eight had impaired fasting glucose (prediabetes) and classified therefore as subjects without T2D alleviation after bariatric surgery. From the total 48 subjects with complete follow-up data, 24 subjects were categorized as good responders (i.e. losing more than 60% of excess BMI within one year), whereas 13 subjects were classified as poor responders (**supplementary table 2**, **supplementary figure 3**). Poor responders not only lost less weight, but also benefited less from bariatric surgery in regards to improvement of lipid metabolism, insulin resistance or inflammation, as they still displayed significantly higher HOMA-IR, triglycerides, LDL-Cholesterol and CRP compared to good responders one year after bariatric surgery (**supplementary table 2**).

**Table 3.** Clinical and biological characteristics of study cohort subjects before, 3 and 12 months after bariatric surgery procedure

|  | Baseline<br>N=48 | Three months post bariatric<br>surgery<br>N=48 | One year post bariatric<br>surgery<br>N=48 |
| --- | --- | --- | --- |
| **General and body composition** |  |  |  |
| **Sex (F/M)** |  | 32/12 |  |
| **Metabolic syndrome status, N (%)** | 36/47 (76.6)<sup>A</sup> | 17/41 (41.5)<sup>B</sup> | 10/43 (23.3)<sup>C</sup> |
| **Diabetes status, N (%)** | 24 (50.0)<sup>A</sup> | 9(18.7)<sup>B</sup> | 4(8.3)<sup>C</sup> |
| **BMI (kg/m<sup>2</sup>)** | 49.7 ± 6.50<sup>A</sup> | 41.3 ± 5.85<sup>B</sup> | 35.1 ± 5.62<sup>C</sup> |
| **WHR** | 0.95 [0.91;1.02]<sup>A</sup> | 0.94 [0.88;0.99]<sup>A</sup> | 0.94 [0.87;1.01]<sup>A</sup> |
| **Total fat mass (in %)** | 49.4 [43.3;53.0]<sup>A</sup> | 46.0 [38.8;48.2]<sup>B</sup> | 37.0 [28.9;44.0]<sup>C</sup> |
| **Visceral fat mass ratio** | 19.0 [17.0;22.0]<sup>A</sup> | 15.0 [12.8;16.0]<sup>B</sup> | 12.0 [8.00;13.0]<sup>C</sup> |
| **Glycemia, insulin resistance and antidiabetic medication intake** |  |  |  |
| **HbA1c (%)** | 5.62 [5.42;6.66]<sup>A</sup> | 5.42 [5.17;5.77]<sup>B</sup> | 5.21 [4.89;5.64]<sup>C</sup> |
| **FPG (mmol/l)** | 5.50 [5.21;7.54]<sup>A</sup> | 5.25 [4.89;6.04]<sup>B</sup> | 5.00 [4.68;5.57]<sup>C</sup> |
| **HOMA-IR** | 5.37 [3.16;8.85]<sup>A</sup> | 2.62 [2.12;2.93]<sup>B</sup> | 2.30 [1.07;3.15]<sup>B</sup> |
| **Hypertension status** |  |  |  |
| **Systolic blood pressure (mmHg)** | 132 ± 12.9<sup>A</sup> | 120 ± 14.9<sup>B</sup> | 120 ± 14.7<sup>B</sup> |
| **Diastolic blood pressure (mmHg)** | 76.6 ± 12.1<sup>A</sup> | 68.2 ± 13.0<sup>A</sup> | 68.8 ± 11.4<sup>A</sup> |
| **Dyslipidemia and antihyperlipidemic medication intake** |  |  |  |
| **LDL-C (mmol/l)** | 3.16 [2.48;3.81]<sup>A</sup> | 2.26 [1.92;2.83]<sup>B</sup> | 2.37 [1.93;2.91]<sup>B</sup> |
| **HDL-C (mmol/l)** | 1.21 [0.99;1.40]<sup>A</sup> | 1.12 [0.96;1.35]<sup>A</sup> | 1.52 [1.27;1.59]<sup>B</sup> |
| **TG (mmol/l)** | 1.68 [1.28;2.15]<sup>A</sup> | 1.15 [0.94;1.46]<sup>B</sup> | 0.96 [0.64;1.30]<sup>C</sup> |
| **Blood and Inflammatory markers** |  |  |  |
| **Leucocytes (Gpt/l)** | 7.50 [6.70;8.50]<sup>A</sup> | 6.60 [5.50;8.03]<sup>A</sup> | 6.10 [5.20;7.32]<sup>A</sup> |
| **LBP (µg/ml)** | 20.465[17.89;24.21]<sup>A</sup> | 15.99 [11.75;17.59]<sup>B</sup> | 6.005 [2.36;14.01]<sup>C</sup> |
| **sCD14 (ng/ml)** | 3089 ± 384<sup>A</sup> | 3626 ± 323<sup>B</sup> | 3243 ± 385<sup>C</sup> |
| **CRP (pg/ml)** | 8.61 [3.87;18.8]<sup>A</sup> | 4.56 [1.49;10.1]<sup>B</sup> | 1.24 [0.43;3.52]<sup>C</sup> |
| **Bacterial DNA amount (in pg per µg extracted whole DNA)** | 0.64 [0.48;1.05]<sup>A</sup> | 0.48 [0.32;0.68]<sup>B</sup> | 1.24 [0.90;2.03]<sup>C</sup> |

Median and first, as well as third, quartile limits (median [q1; q3]) are shown for non-normally distributed, continuous variables. For normally distributed, continuous variables, data are given in mean ± standard deviation (mean ± SD). For categorical parameters, total numbers (percentage) are shown. Significant p-values are depicted according to group differences: the difference between 2 values in a row with the same letter are non-significant. **Abbreviations:** **BMI:** Body mass index; **CRP:** C-reactive protein; **FPG:** Fasting plasma glucose; **HbA1c:** Glycated Hemoglobin A1c; **HDL:** High density lipoprotein; **HOMA-IR:** Homeostatic model assessment for insulin resistance; **LBP:** Lipopolysaccharide binding protein; **LDL:** Low density lipoprotein; **sCD14:** soluble CD14; **TG:** Triglycerides; **WHR:** Waist to hip ratio.

### Reduced bacterial blood load in T2D is ameliorated after bariatric surgery

Bacterial DNA could be measured in 127 blood samples out of 144 samples with available follow-up data at all three timepoints at a quantity ranging from 4.63 to 140.4 fg in 200 µl blood (median=21.6 fg). Albeit using the same procedure and the same kit, total extracted DNA varied widely between samples spanning from 12.7 to 98.2 ng total DNA/µl blood leading to a corrected range of bacterial DNA quantity within 0.08 - 4.11 pg per µg total DNA (median= 0.7 pg bacterial DNA per µg total DNA). While bacterial DNA could be detected in negative controls at 0.162 pg per total g DNA, total DNA in negative controls did not exceed 1.2 ng/µl leading to total bacterial DNA amounts of less than 1.5 fg (median= 0.4 fg). Overall, bacterial blood load was positively associated with bacterial richness at baseline (**fig 4A**) and negatively with Leukocytes (**fig 4B**). This observation was further validated in a replication cohort consisting of 62 subjects from the same center (**supplementary table 3**). Correlation analyses in the replication cohort further underline the negative associations between inflammation and bacterial load and the association between LBP and inflammation as well as obesity (**supplementary table 6**).

**Figure 4.**
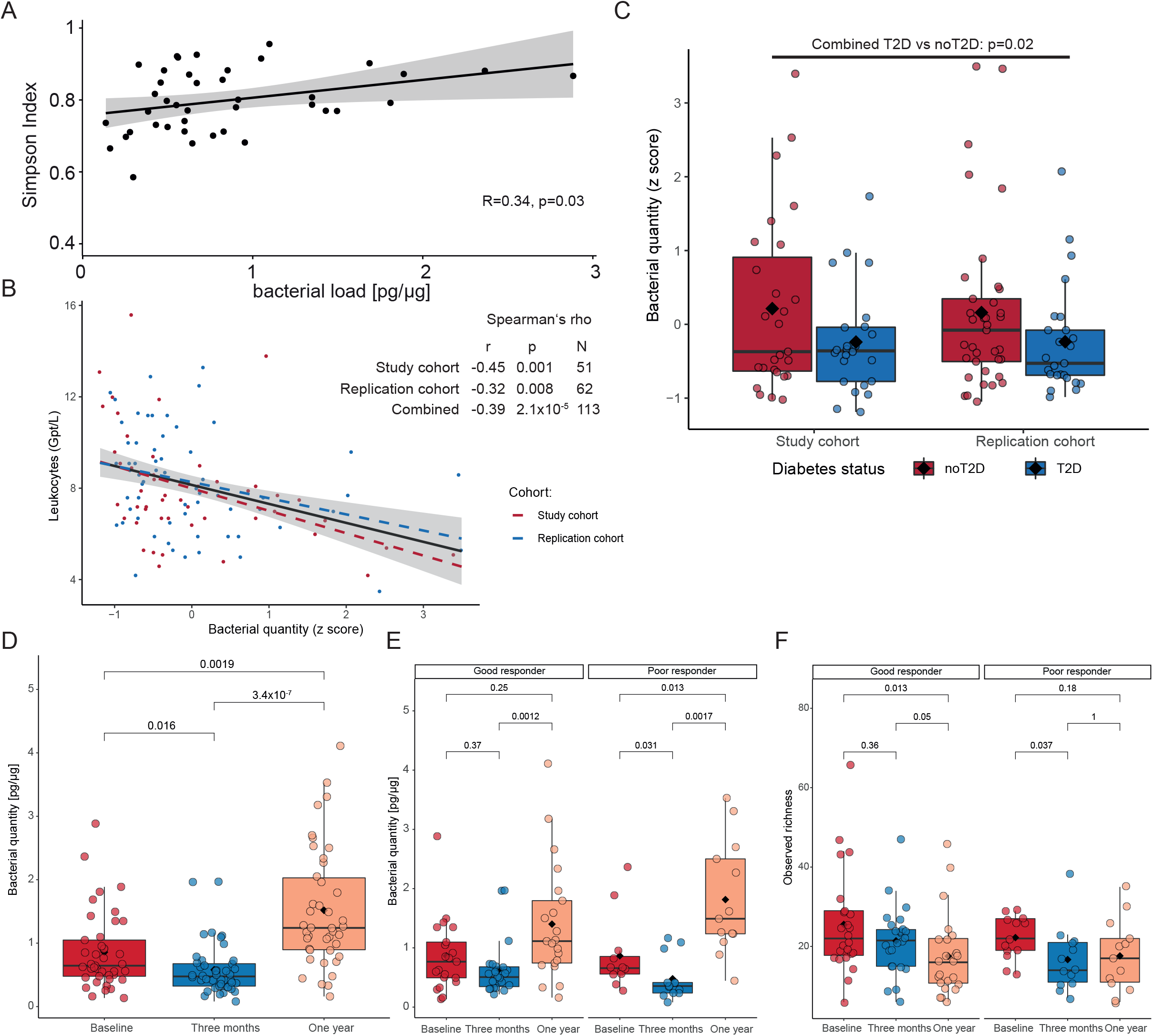
**(A)** Spearman’s rank correlation of bacterial load at baseline with alpha diversity (Simpson Index) the grey area around regression line indicates the confidence interval at a confidence level of 95%. **(B)** Spearman’s rank correlation of bacterial load at baseline with Leukocytes in the study and replication cohort. Colors depict cohorts with red being specific to the study cohort and blue to the replication cohort. Bacterial quantity is standardized within cohorts. The grey area around regression line indicates the confidence interval at a confidence level of 95%. **(C)** Blood bacterial in load in T2D vs nonT2D in both study and replication cohort. Bacterial quantity was standardized prior to statistical analysis. Boxplots are shown with Tukey-whiskers and mean (◆) as well as median. T2D was compared with nonT2D using Wilcoxon signed rank test after pooling both cohorts. **(D)** Bacterial quantity in pg per µg extracted DNA over time. The three timepoints are compared using Kruskal-Wallis test and results are validated via Friedman’s test (Kruskal-Wallis p-value is depicted). Paired samples Wilcoxon signed-rank test is used to compare two groups at once. Only samples with available triplicates in blood bacterial load are used for the comparisons (n=39). **(E)**-Bacterial quantity dynamics in good and poor responders over time (n=21 good responders, n=13 poor responders). The three timepoints are compared using Kruskal-Wallis test and results are validated via Friedman’s test (Kruskal-Wallis p-value is depicted). Paired samples Wilcoxon signed-rank test is used to compare two groups at once. **(F)** Diversity dynamics in good and poor responders over time (n=24 good responders, n=13 poor responders). The three timepoints are compared using Kruskal-Wallis test and results are validated via Friedman’s test (Kruskal-Wallis p-value is depicted). Paired samples Wilcoxon signed-rank test is used to compare two groups at once.

Spanning over all timepoints, bacterial load was negatively correlated with waist circumference, HbA1c, and inflammation markers such as leukocytes as well as markers related to a presumed leaky gut such as LBP (**supplementary fig 6A-D, supplementary tables 7 and 8**). The latter was on the other hand associated with markers of metabolic disease and obesity (**supplementary table 7**). While these correlations were weak to moderate, the use of Spearman’s rank correlation made them less likely to be influenced by a few high leverage points as shown after removing statistical outliers for bacterial quantity (**supplementary figure 7A-D**). Accordingly, subjects with T2D at baseline displayed lower bacterial quantity in both our study and replication cohorts (**fig 4C**).

Blood bacterial load was decreased three months post bariatric surgery (median bacterial quantity in pg/µg at baseline = 0.032, vs 0.024 at three months, p=0.013, **fig 4D**), but increased significantly at one year following surgery (median bacterial quantity in pg/µg at one year = 0.062, p-value compared to three months=1.1×10^-6^, p-value compared to baseline =0.002, **fig 4D)** independently of T2D status at baseline **(supplementary figure 8A, B).** When checking the changes according to weight loss response, subjects deemed ‘good responders’ and ‘poor responders’ displayed similar blood bacterial load at baseline (median bacterial quantity in pg/µg ‘good responder’ = 0.864 vs ‘poor responder’ = 0.866, p-value = 0.9) but blood bacterial quantity at one year increased less dramatically in good responders and was not significant as compared the increase in blood bacterial quantity in poor responders (fold change 1.6 vs 2.1 in good vs poor responders respectively, **fig 4E** ). Bacterial diversity on the hand, showed a transient significant decrease at three months only to increase again almost to baseline at one year in poor responders, while continuously and significantly decreasing in good responders (**fig 4F**).

LBP, expected to reflect host’s response to bacterial DNA, decreased continuously and significantly after bariatric surgery (median LBP in µg/µl at baseline = 20.47 vs 15.99 at three months and 6.0 at one year after bariatric surgery, p_3MonthvsBaseline_ = 3.3e^-5^, p_1Yearvs3Months_ = 2.46e^-6^ p_1YearvsBaseline_ =5.8e^-9^) independently of weight loss response or diabetes status at baseline (**supplementary figures 9A, B**). Similarly, leukocytes decreased overtime after bariatric surgery but less so and non-significantly in subjects with poor response (good responders: p_3MonthvsBaseline_ = 3.3e^-5^, p_1Yearvs3Months_ = 0.14 and p_1YearvsBaseline_= 6.5e^-4^ respectively; poor responders: p_3MonthvsBaseline_ = 0.67, p_1Yearvs3Months_ = 0.08 and p_1YearvsBaseline_ = 0.16) (**supplementary fig 9C**).

### Metabolic improvement after bariatric surgery is accompanied by early changes in microbial differential abundances in subjects with and without T2D

Beyond changes in blood bacterial load over time, there were significant and consistent shifts in bacterial genera over the whole cohort between month three and twelve after bariatric surgery as compared to baseline. Decreased ASVs belonged to genera such as *Anoxybacillus*, *Rhizobacter* and *Sphingomonas*, whereas other genera such as *Acinetobacter, Granulicatella* and *Pseudomonas* increased. Comparing shifts at one year and baseline showed that most of these seen at three months post bariatric surgery were preserved at one year (**fig 5A**).

**Figure 5.**
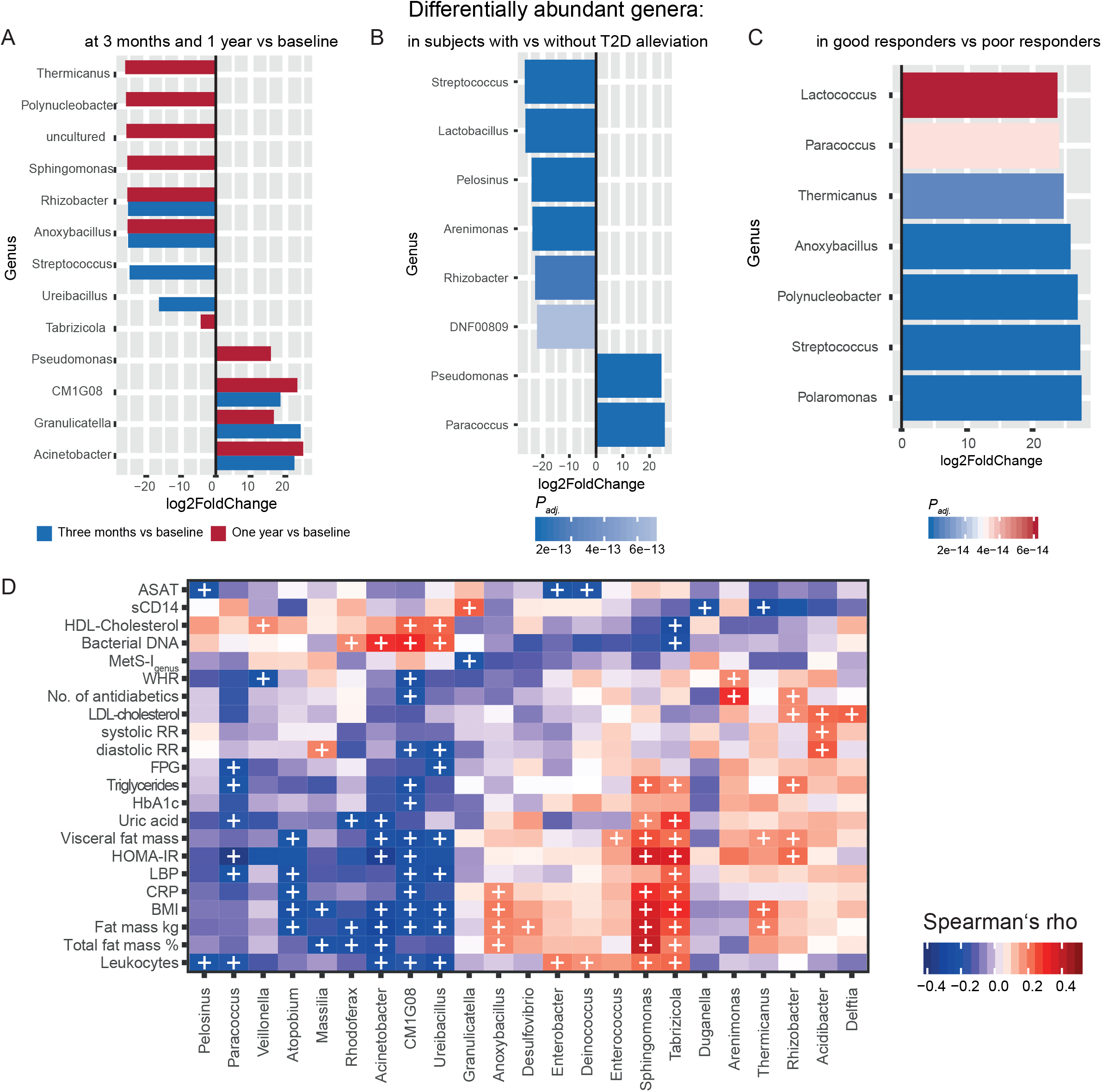
**(A)** Differentially abundant genera between baseline and 3 months (blue bars) and baseline and one year (red) within the complete cohort with available taxonomy at all timepoints (n=48). Differential abundance is calculated on ASV level abundances using taxonomy after stringent control for decontamination at a decontam score 0.5 and reported at genus level. Only significant differential abundances (*Padj* < 0.01) are reported. **(B)** Differentially abundant genera in subjects with T2D, who experienced diabetes remission as compared to those with persistent T2D (n=12 in both groups).Color shading represents p-values. **(C)** Differentially abundant genera in good responders vs poor responders (n=24 vs 13 respectively). Color shading represents p-values. **(D)** Spearman’s rank correlations of differentially abundant genera in good responders and subjects with T2D remission with host variables in all timepoints. (+) indicates a significance level <0.05.

Moreover, T2D remission following bariatric surgery was associated with significantly lower proportions of *Burkholderiacae*, *Veilonellaceae* and *Lactobacillaceae* and higher proportions of *Rhodobacteraceae* and *Pseudomonales* at all timepoints combined **(fig 5B)**. Genera with observed shifts after bariatric surgery were closely tied to metabolic control: Genera, which had a lower abundance after surgery such as *Anoxybacillus, Rhizobacter* and *Sphingomonas* e.g. were significantly positively associated with markers of body composition such as. overall and visceral fat mass, BMI and CRP. Furthermore, significantly positive correlations were observed for aforementioned genera with markers of metabolic control including HOMA-IR, number of antidiabetics, uric acid, triglycerides and LDL-Cholesterol (**fig 5D**). As for genera being more abundant after surgery, C*M1G08* and *Acinetobacter* were negatively associated with markers of central obesity and metabolic disease as well as inflammation, while being positively associated with HDL-cholesterol and blood bacterial quantity. Contrarily, *Granulicatella* was negatively associated with excess BMI loss, but positively with sCD14. Of note, while *Granulicatella* was higher at one year compared to baseline, it seemed to peak at three months, while the increase in *GM1G08* and *Acinetobacter* was continuous (**fig 5A**). *Paracoccus –* which was shown to be decreased in T2D at baseline and increased in subjects experiencing T2D remission - was negatively associated with inflammation, fasting glucose, triglycerides, HOMA-IR and LBP (**fig 5D**).

Good responders displayed an increase in ASVs belonging to *Streptococcaceae*, such as *Lactococcus* and *streptococcus* species, *Burkholderiales* such as *Polaromonas* and *Polynucleobacter*, *Rhodobacteraceae* such as *Paracoccus* and *Bacillaceae* such as *Anoxybacillus* at all timepoints (**fig 5C**). While *Paracoccus* was associated with decreased inflammation and metabolic impairment, some genera increased in good vs poor response were associated with increased BMI and inflammation such as *Anoxybacillus* or *Thermicanus* overall, but significantly decreased at one year when compared to baseline (**fig.5A**) Several genera abundantly observed in good responders and subjects who experience T2D remission were initially decreased in T2D, further delineating a putative positive association with good metabolic health (**fig 3B, 5B-D**).

### CARD-FISH in blood the visualization of living bacteria post bariatric surgery

In order to verify whether increased bacterial quantity post bariatric surgery is only related to uptake of bacterial sequences or bacteremia, we sought to visualize bacterial cells in a subject, who had undergone bariatric surgery, in whom blood bacterial quantity could be measured at baseline and for whom bacterial sequence data are available. CARD-FISH implementation was unsuccessful in frozen blood samples, but considering blood bacterial quantity increased after surgery over time, we hypothesized that this state is more likely to yield bacterial cell visualization. Because bacteremia and endotoxemia have been associated with chewing and food intake [5, 54, 55], blood samples were also collected 1 h after a mixed meal to increase chances of visualization. Similarly, samples were taken from a healthy athletic control, whose weight has been stable for the last 2 years and who has no known diseases and no medication.

Our CARD-FISH results show the presence of bacteria in the blood samples from the patient collected before and after mixed meal intake with an abundance of 7.9 × 10^4^ cells mL^-1^ and, 1.2 x 10^5^ cell mL^-1^ respectively (**supplementary figure 10 A,B, Supplementary file 8**) in support of increased bacteremia after food intake and chewing. In contrast, blood samples from the healthy control reveled no presence of hybridized bacteria, although filter pieces from all filters (3 µm and 0.2 µm) used for cell separation of both the supernatant and Nycodenz layers were hybridized and imaged (supplementary figure 10 C,**D)**. An additional control to certify the CARD-FISH was successful and the lack of hybridized cells in the healthy subject is not due to a technical error, same control blood sample was deliberately infected with *Pseudomonas putida* (DSM6125) (**supplementary figure. 10 E,F**). Therein, positively hybridized *P. putida* cells could be observed and no other cell morphotypes, adding evidence that freshly collected blood samples from the healthy control do not contain bacteria at abundances that can be detected by CARD-FISH.

## Discussion

The presence of bacteria and bacterial products in various tissues and their contributions to the local and systemic inflammation have been suggested as novel mechanisms for both development and progression of “non-communicable” metabolic diseases, such as obesity [13–15, 17, 19, 22], T2D [13–15], cardiovascular diseases [17] and cirrhosis [18, 56]. However, studies supporting this hypothesis are mostly limited by the lack of control for contamination and a high overestimation of bacterial DNA quantity, consequently inflating results. In the present work, we therefore quantified and characterized bacterial DNA in the blood of subjects with high metabolic burden undergoing bariatric surgery with an emphasis on including and subtracting suitable negative controls. The goal was to identify links between metabolism and a putative blood bacterial signature as well as its dynamics over time after bariatric surgery.

After stringent experimental and bioinformatic control for contaminants, we report 2746 features (ASVs) belonging mostly to the phyla *Proteobacteria* and *Firmicutes*, which corroborates previously published results [13, 14, 21, 56, 57]. Furthermore, we evidence quantitative, compositional and taxonomic signatures associated with markers of metabolic disease and T2D and shifts thereof driven by medication and weight loss intervention. We also provide qualitative and quantitative evidence for the presence of bacterial cells in the blood samples of a subject with obesity, who has undergone bariatric surgery as well as bacteremia after mixed meal intake in comparison to a healthy control.

Our results provide support for the existence of an individually determined core circulatory bacterial signature and microbiome, which reflects disease and intervention with 40% of observed variance in bacterial composition explained by 27 host variables prominently related to inflammation, anthropometric measures as well as metabolic health and medication intake. Of the latter, notable associations of bacterial signature were seen for metformin and PPI intake. These results underscore findings from other studies relying on the application of multi-method characterizations including evidence of viability to debunk the notion that the blood is free of viable microorganisms [14, 56–61] as well as studies showing the deeply underrated modulatory effect of medication on host microbial communities and the importance of assessing drug intake in microbial surveys of any niche [62].

Moreover, blood bacteria signature allowed the classification of subjects with and without MetS along their actual clinical group and more accurately so at genus than phylum level. This is in line with recent evidence showing disease specific tissue signatures in several tissues pertaining to metabolic health as well as cancer [13, 14, 21] and could be related to the particular vulnerability to translocation of specific ingested bacteria in respective diseases. Together, these independent observations reinforce the notion of a predictive circulatory metabolic bacterial signature reminiscent of the described enterotypes [63] in the gut microbiome.

Similarly, we show a specific bacterial signature for T2D characterized by reduced bacterial diversity and an overall reduction in genera belonging to *Firmicutes* and *Proteobacteria*. Diversity scores were negatively correlated with inflammation, visceral adiposity and uremia, echoing observations of diversity in gut microbial communities in T2D [64]. Although the reported reduction in genera is in line with data from Anhê et al., we did not evidence their reported reduction in *Bacteroidetes* nor the increase in *E.coli* in T2D [13]. While we evidence ASVs belonging to *Bacteroidetes*, their prevalence was very low and was not particularly associated with metabolic control in our cohort. This could be indeed due either to cohort specific characteristics or our choice in primers. Our data are more in accordance with results from another cohort based in our center. On the other hand, we report several more genera in the blood (314 with 120 core genera) compared, which possibly reflects our choice of whole blood as material compared to plasma samples used by Anhê et al.

A hallmark for T2D in our study was the loss of genera related to *Bacillaceae* and *Bukholderiaceae*. Several of the reduced genera were associated with a healthier metabolic status. Others, such as *Sphingomonas* and *Acidibacter* were positively associated with BMI and insulin resistance as well as increased blood pressure and antihyperlipidemic medication respectively. Considering subjects with and without T2D at baseline were matched on BMI and did not differ significantly in blood pressure nor lipid levels but were significantly more medicated, these differences could arise from medication effects. This is supported by evidence from Anhê et al. showing increased *Sphingomonas* in subjects with T2D (at least in Adipose tissue) and by the fact that these differences disappear after bariatric surgery in our cohort over time, when subjects are taken off their antihyperlipidemic and antihypertensive medication sequentially.

Bariatric surgery, on the other hand, led to swift changes in metabolism and bacterial composition in the blood: Changes occurring after bariatric surgery take place early on and are mostly stable over time, which is in line with available data on the rearrangement of the gut microbiome after bariatric surgery [65]. In addition, subjects who experienced T2D remission showed a significant decrease in several clades associated with cardiometabolic and cardiovascular disease such as *Streptococcaceae* [66], *Veilonellaceae* [67] or *Lactobacillales* [68] which are evidenced to be increased in T2D and under diabetic medication in the gut. In contrast, a few health-related genera increased after bariatric surgery, i.e. genera associated with improved insulin sensitivity and ameliorated lipid status or reduced adiposity at baseline in our cohort. Similarly, subjects who experience T2D remission after bariatric surgery displayed increases in genera seen to be initially decreased in T2D, further delineating a positive putative association of these bacteria with metabolic health.

We further investigated circulating bacterial DNA load: Quantification of 16s rRNA showed that bacterial load was positively associated with bacterial diversity and that the increase of the very small bacterial quantity observed after bariatric surgery reflects a reduced inflammatory tone and an improved metabolic health, which we found intriguing. This was also supported by the observations that a) bacterial load is reduced in T2D at baseline, also in our independently measured replication cohort, b) bacterial load was negatively associated with metabolic disease index score at baseline, c) bacterial load was associated with bacterial composition in the opposite direction dictated by metabolic risk factors such as HbA1c, BMI and TG and that d) it increased after bariatric surgery. This is further corroborated by the fact that all associations with bacterial quantity are positive with health markers at each and all timepoints. There was no significant difference in bacterial load at baseline between good and poor responders but poor responders experienced an even more pronounced increase in bacterial load. Unexpectedly, bacterial diversity continuously and significantly decreased in good responders, while in poor responders it significantly dipped around 3 months only to almost recover at one year post bariatric surgery. This was paralleled by similar dynamics for leukocytes in good responders, while poor responders failed to display a significant decrease in Leukocytes and inflammation over time. Considering bacterial diversity was still negatively associated with leukocytes at all timepoints, it seems likely that the superior increase in bacterial quantity in poor responders is the driving factor behind the observed increased diversity. The latter, on the other hand and along increased bacterial quantity, could be at least partially responsible for the sustained inflammation seen at one year in poor responders. Thus, the increased bacterial quantity after bariatric surgery could be related to increases in gut permeability [69] and transmissibility of oral bacteria after the surgery due to increased pH [70]. Stronger increases thereof in poor responders are possibly related to an even more impaired gut permeability due to sustained obesity [71] as well as increased food intake. This begs the question of the transient reduction in bacterial quantity at three months post bariatric surgery: Considering the origins of bacterial DNA we see is very likely environmental (i.e. food and oral bacteria), bacterial quantity reduction at three months could be related to the extreme restriction of food intake, far more pronounced at three months than around one year after surgery where eating behavior and weight loss are more stable. In support of this, bacterial diversity decreases at three months in both good and poor responders only to increase in poor responders again, which could indeed be due to increased food intake. It is important to note that the observations are based on an increase in miniscule amounts of bacterial DNA far from the expected quantity in clinically relevant bacteremia and sepsis.

We would like to acknowledge limitations in our study: Although we control for contamination starting at DNA isolation including filling EDTA vials with PBS via needle butterflies to take production line contamination of medical products into account, we cannot fully exclude contamination via puncture of the skin or other environmental sources. Of note, highest diligence was used in skin decontamination prior to venipuncture and DNA isolation vials were collected as the last vials (in a sequence of 12 vials). Moreover, we did not observe typical skin bacteria among identified contaminants, making the skin as a source of contamination unlikely. Spurious environmental contamination cannot be fully excluded and might explain the encounter of *Choloroflexi*, *Rhizobacter*, *Limnohabitans* and *Plactomycetes* as well as *uncultured diatom* in our dataset. While we aimed to reduce these spurious findings as much as possible in a data driven manner to avoid arbitrary preselection of taxa, even complete subtraction of contaminants AVSs selected via stringent decontam score as done by Poore et al. [21] did not eliminate these taxa from the dataset completely. This commends the development of even better pipelines and bioinformatics tools for decontamination in small bacterial biomass samples. An encouraging observation nonetheless, is that these taxa do not seem to be particularly relevant for our observed phenotype associations nor does a more stringent decontamination score change the conclusion of our work, further supporting the emergence of disease signatures in circulating bacterial composition.

We evidence circulatory bacterial DNA but can only speculate about its origins: The translocation of bacteria from the gut has been the primary considered mechanism. Although this hypothesis is tempting, we did not validate it by performing gold standard gut permeability tests. Moreover, evidence from the HMP have shown that the blood bacterial signature closely resembles that from the skin and oral cavity [58], further pointing to alternative or additive origins. Considering transmission of oral strains along the gastrointestinal tract is more common than previously recognized [72–74] and that gut rearrangement after bariatric surgery is associated with increased pH in the gut, a shift in bile acid pools [70] and increased PPI prescription especially after RYGB also evident in our cohort [75], we cannot exclude that the change in the circulatory blood bacterial signature is reflective of changes in these, among several other bacterial host niches. The increase in blood bacterial load after bariatric surgery could be seen as underpinning this hypothesis. While our data make the case, that other mechanisms override an initial increase in gut permeability, which has been associated with insulin resistance and obesity, to induce weight loss and improvement of glucose tolerance after bariatric surgery, we are unable to validate this hypothesis at this point.

Finally, the reported correlations on bacterial quantity and those related to bacterial composition with low effect size should be considered with caution. Despite their statistical significance, the biological relevance of these associations might be questionable and commends further independent replications.

Beyond these restrictions and while the evidence for the presence of bacteria in the placenta has been highly controversial, with independent groups refuting these findings, there has been no evidence in the literature that convincingly shows that the blood is a tissue constantly free from bacteria or bacterial genetic material. The limitations of the several studies linking bacteria in the blood with disease have been addressed in the present work further adding to its strengths: Diligent sterile handling of materials and pre-treatment of lab materials with UV-light was employed. We further included several negative controls (PBS for extraction and blank controls for PCR), while further accounting for production line contamination of medical products using the same tubes and medical materials used to draw the blood in our subjects. These negative controls were confirmed in agarose gel but were still sequenced alongside the true samples. Moreover they were actively used to identify possible contaminants in the dataset which we then excluded using the same Software, that had helped debunk or at least shake the pertinacious notion of a ‘placenta microbiome’ [76, 77]. We further replicated findings on bacterial load in an independent cohort, which was processed and analyzed independently. Moreover, we could substantiate the associations between host metabolic health with individual taxa by several different statistical methods while accounting for the impact of relevant covariates, which has, to our knowledge, never been done performed. Beyond a mere diagnostic tool for metabolic disease, which can be currently achieved more easily and at a lower cost, the current work underpins the notion that the blood is a putative ecological niche. Despite extensive immune control it remains dynamic and reflects metabolic health, warranting further contaminant conscious rigorous work to delineate mechanistic and exploitable pathways and targets similar to the application of pasteurized bacteria (e.g. *Akkermansia muciniphila*) [78–83] for weight loss and metabolic improvement.

## Conclusions

In summary, while reducing and controlling both for contamination and technical biases, we could detect low amounts of bacterial DNA in the blood, where we are able to derive a metabolic fingerprint reflected in bacterial DNA composition. We could also observe differences in diversity, amount of bacterial DNA and bacterial composition between subjects with and without T2D, subjects with or without T2D remission or with varying degrees of weight loss response to bariatric surgery. We further substantiate our findings of bacterial quantity increase after bariatric surgery with imaging of live bacteria in post bariatric patients. The present work lays a stepping stone and encourages rigorous cross sectional and longitudinal studies in larger cohorts with both diseased and healthy subjects. These studies should optimally expand over various expertise to provide insights into the functionality and potential role of a ‘circulatory microbiome’ in maintaining health and contributing to the onset and progression of disease.

## Data Availability

The datasets as well as code analyzed and used during the current study are available under reserved DOIs and will be published after acceptance (citation will be incorporated).
Raw data is available under individual and reasonable request to the authors.

## Abbreviations

ALAT: Alanin-Aminotransferase
ARBs: Angiotensin II receptor blockers
ASAT: Aspartat-Aminotransferase
BMI: Body mass index
CRP: C-reactive protein
EBL: Excess BMI loss
eGFR: Estimated glomerular filtration rate
FPG: Fasting plasma glucose
GLP1: Glukagon-like peptide 1
HbA1c: Glycated Hemoglobin A1c
HDL: High density lipoprotein;
HOMA-IR: Homeostatic model assessment for insulin resistance
LBP: Lipopolysaccharide binding protein
LDL: Low density lipoprotein
MetS: Metabolic syndrome
NGT: Normal glucose tolerance
PPI: Proton pump inhibitors
RDA: Redundancy analysis
rRNA: ribosomal ribonucleic acid
sCD14: soluble cluster of differentiation 14
T2D: Type 2 diabetes
WHR: Waist to hip ratio

## Declarations

### Ethics approval and consent to participate

This study was approved by the ethics committee of the University of Leipzig (application number: 047- 13-28012013) and all participants involved gave written informed consent. The research was performed in accordance with the principles of the Declaration of Helsinki.

### Consent for publication

Not applicable

### Availability of data and materials

The datasets as well as code analyzed and used during the current study are available under reserved DOIs and will be published after acceptance (citation will be incorporated).

### Competing interests

The authors have no conflict of interest to declare.

### Funding

This work was supported by grants from the Deutsche Forschungsgemeinschaft (DFG, German Research Foundation – Projektnummer 209933838 – SFB 1052; B01, B03, B09) and from IFB AdiposityDiseases (AD2-060E, AD2-06E95, and AD2-K7-117). IFB Adiposity Diseases is supported by the Federal Ministry of Education and Research (BMBF), Germany, FKZ: 01EO1501. RC was supported by a junior research grant by the Medical Faculty, University of Leipzig and by the Federal Ministry of Education and Research (BMBF), Germany, FKZ: 01EO1501 (IFB AdiposityDiseases, MetaRot program). The content of this work is the responsibility of the authors and does not represent the official views of the funding agencies.

### Author contributions

RC and PK designed the study; RC performed experiments; RC and LM analyzed the data; NM supervised and conducted CARD-FISH experiments and analyzed related downstream data. NS performed quantification of bacterial cells after CARD-FISH. All authors contributed to manuscript preparation and writing.

## Acknowledgment

We would like to thank Stefanie Walther, Judith Kammer, Charlott Krahnepuhl and Eileen Bösenberg for their excellent support in recruiting subjects, supervising subjects during the study and aftercare, organization and coordination of the studies, and sample, as well as diligent data collection. We also thank Stefanie Ziesche for excellent technical assistance in the lab. We would further like to thank all subjects who took part in the study and without whom this research would not have been possible. We acknowledge the Centre for Chemical Microscopy (ProVIS) at the Helmholtz Centre for Environmental Research for using their sample preparation and microscopy facilities. Special thanks to Katja Nerlich for support during in situ hybridization and fluorescence microscopy image acquisition. **Supplementary figures 1 and 3** were created using BioRender.com.

## Supplementary Material

### Supplementary Tables

**Supplementary Table 1.**
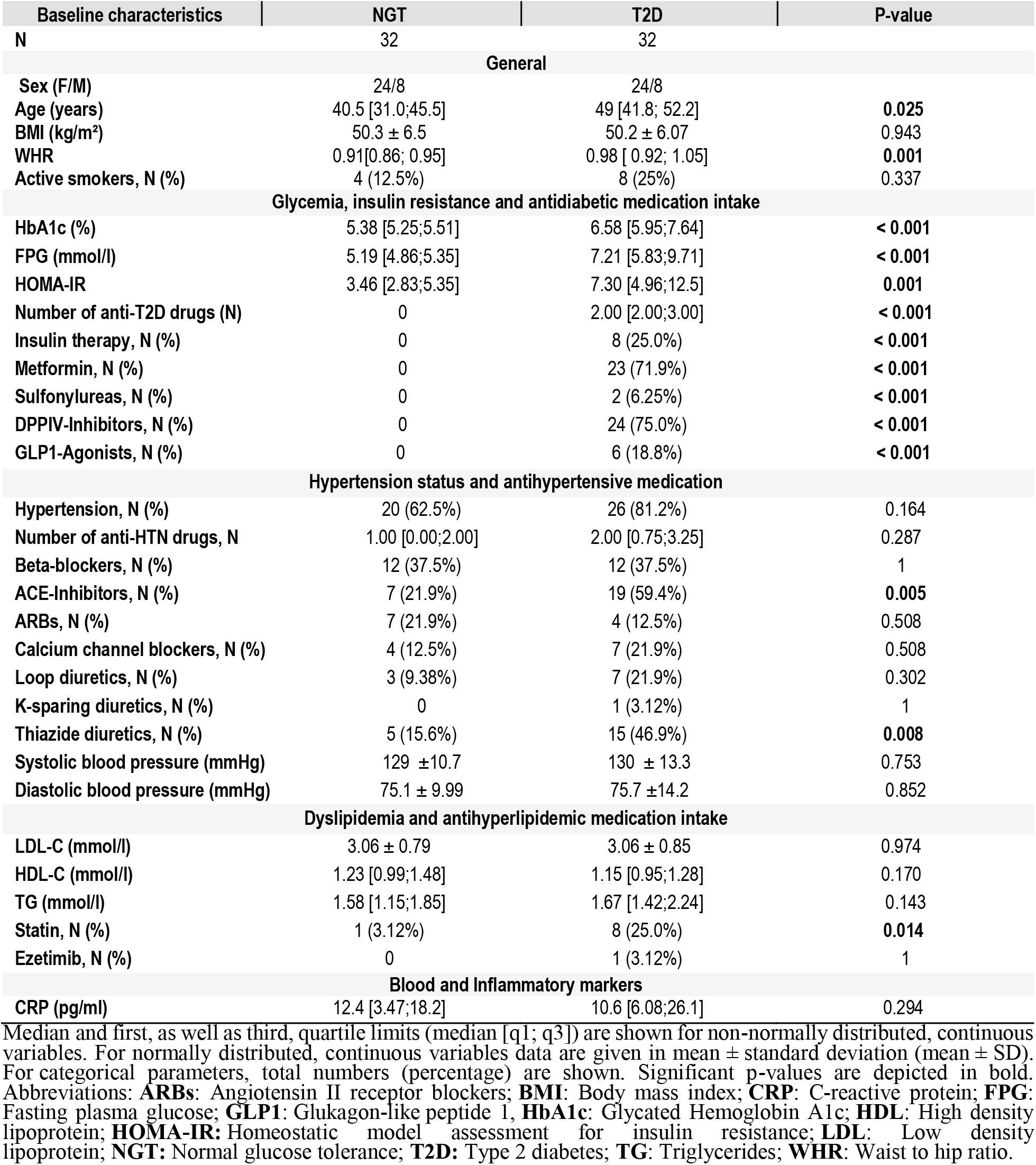
baseline cohort characteristics with initial matching (n=64)

**Supplementary Table 2.** Phenotypes of good vs poor responders at baseline and one year post bariatric surgery

| Baseline characteristics | Good Responder |  | Poor Responder |  | P-value |  |
| --- | --- | --- | --- | --- | --- | --- |
|  | baseline | One year | baseline | One year | baselines | One year |
| N | 24 |  | 13 |  |  |  |
|  | General |  |  |  |  |  |
| Sex (F/M) | 17/7 |  | 10/3 |  |  |  |
| Age (years) | 46.3 ± 8.87 | 46.3 ± 8.87 | 48.2 ± 6.14 | 48.2 ± 6.14 | 0.79 |  |
| BMI (kg/m²) | 47.7 ± 5.59 | 30.8 2.97 | 51.7 ± 6.58 | 40.9 ± 3.39 | 0.14 |  |
| WHR | 0.96 ± 0.11 | 0.91 [0.86;0.98] | 0.93 ± 0.06 | 0.94 [0.90;0.96] | 0.82 |  |
| Visceral fat ratio | 18.8 ± 4.69 | 8.00 [6.00;9.25] | 21.2 ± 5.97 | 13.0 [13.0;15.0] | 0.46 |  |
|  | Glycemia, insulin resistance and metabolic syndrome at baseline |  |  |  |  |  |
| T2D Status (Yes) | 11 (45.8%) | 2 (8.33%) | 9 (69.2%) | 2 (15.4%) | 0.46 | 0.6 |
| HbA1c (%) | 6.56 ± 1.70 | 5.13 [4.80;5.44] | 6.24 ± 1.16 | 5.41 [4.94;5.96] | 0.78 | 0.24 |
| FPG (mmol/l) | 7.28 ± 3.46 | 5.01 [4.66;5.57] | 7.22 ± 3.07 | 5.33 [4.78;5.81] | 1 | 0.44 |
| HOMA-IR | 13.8 ± 25.9 | 1.24 [0.77;2.72] | 7.00 ± 7.55 | 3.20 [2.53;3.53] | 0.63 | **0.02** |
| Antidiabetis N | 9 (37.5%) | 2 (8.33%) | 9 (69.2%) | 2 (15.4%) | 0.27 | 0.60 |
| MetS Status | 19 (79.2%) | 4 (18.2%) | 9 (69.2%) | 4 (33.3%) | 1 | 0.60 |
|  | Hypertension status and antihypertensive medication |  |  |  |  |  |
| Systolic RR (mmHg) | 129 ± 13.5 | 118 ± 15.9 | 137 ± 12.5 | 124 ± 13.2 | 0.30 | 0.23 |
| Diastolic RR (mmHg) | 76.0 ± 12.3 | 67.3 ± 10.7 | 76.0 ± 11.8 | 69.9 ± 11.7 | 1 | 0.5 |
|  | Dyslipidemia and antihyperlipidemic medication intake |  |  |  |  |  |
| LDL-C (mmol/l) | 3.18 ± 1.04 | 2.08 [1.83;2.68] | 3.24 ± 0.69 | 2.67 [2.42;2.97] | 0.98 | **0.03** |
| HDL-C (mmol/l) | 1.26 ± 0.40 | 1.54 [1.33;1.74] | 1.25 ± 0.25 | 1.52 [1.35;1.55] | 0.99 | 0.44 |
| TG (mmol/l) | 1.84 ± 0.84 | 0.86 [0.62;1.08] | 1.75 ± 0.59 | 1.21 [1.01;1.50] | 0.93 | **0.03** |
| Statin, N (%) | 4 (16.0%) | 4 (16.7%) | 3 (23.1%) | 3 (23.1%) | 0.96 | 0.67 |
|  | Blood and Inflammatory markers |  |  |  |  |  |
| Haemoglobin (g/dl) | 13.9 ± 1.18 | 13.1 ± 1.21 | 13.7 ± 0.64 | 13.0 ± 1.21 | 0.88 | 0.83 |
| Leucocytes (Gpt/l) | 7.99 ± 2.45 | 6.23 ± 1.58 | 7.65 ± 1.75 | 6.84 ± 1.89 | 0.88 | 0.33 |
| CRP (pg/ml) | 11.7 ± 9.86 | 0.94 [0.38;1.60] | 14 ± 11.8 | 3.41 [1.12;5.36] | 0.95 | **0.018** |
| Bacterial load fg/ng | 0.86 ± 0.62 | 1.11 [0.74;1.80] | 0.87 ± 0.62 | 1.49 [1.24;2.50] | 1 | 0.14 |

Continuous variables data are given in mean ± standard deviation. Median and first, as well as third, quartile limits (median [q1; q3]) are shown for non-normally distributed. For categorical parameters, total numbers (percentage) are shown. Significant p-values are depicted in bold. Abbreviations: **BMI**: Body mass index; **CRP**: C-reactive protein; **FPG**: Fasting plasma glucose; **HbA1c**: Glycated Hemoglobin A1c; **HDL**: High density lipoprotein; **HOMA-IR**: Homeostatic model assessment for insulin resistance; **LDL**: Low density lipoprotein; **NGT**: Normal glucose tolerance; **RR**: Blood pressure (Riva Rocci) **T2D**: Type 2 diabetes; **TG**: Triglycerides; **WHR**: Waist to hip ratio.

**Supplementary Table 3.** Replication Cohort characteristics

| Baseline characteristics | NGT | T2D | P-value |
| --- | --- | --- | --- |
| N | 37 | 25 |  |
| **General** |  |  |  |
| Sex (F/M) | 27/10 | 16/9 | 0.57 |
| Age (years) | 44 ± 11 | 53 ± 11 | **0.003** |
| BMI (kg/m<sup>2</sup>) | 46.28 ± 5.63 | 48.46 ± 7.4 | 0.38 |
| **Glycemia, insulin resistance and lipid metabolism** |  |  |  |
| FPG (mmol/l) | 5.41 ± 1.07 | 7.06 ± 1.93 | **0.001** |
| HOMA-IR | 4.70 ± 3.91 | 9.23 ± 12.68 | 0.11 |
| Triglycerides (mmol/l) | 1.28 ± 0.76 | 2.25 ± 2.22 | 0.05 |
| HDL-Cholesterol (mmol/l) | 1.16 ± 0.34 | 0.98 ± 0.28 | **0.04** |
| LDL-Cholesterol (mmol/l) | 2.84 ± 0.72 | 2.57 ± 0.86 | 0.2 |
| **Inflammatory markers** |  |  |  |
| Leukocytes (Gpt) | 8.08 ± 2.24 | 8.54 ± 2.33 | 0.44 |
| LBP | 11.72 ± 4.00 | 14.33 ± 6.26 | 0.07 |

Data are given in mean ± standard deviation (mean ± SD). For categorical parameters, total numbers (percentage) are shown. Significant p-values are depicted in bold. **Abbreviations:** **BMI:** Body mass index; **CRP:** C-reactive protein; **FPG:** Fasting plasma glucose; **HbA1c:** Glycated Hemoglobin A1c; **HOMA-IR:** Homeostatic model assessment for insulin resistance; **LBP:** Lipopolysaccharide binding protein

**Supplementary Table 4:** Envfit output for 27 variables on genera level RDA

|  | RDA1 | RDA2 | R<sup>2</sup> | Adj. P(r) |
| --- | --- | --- | --- | --- |
| Variables |  |  |  |  |
| BMI | 1.00000 | 0.00007 | 0.2219 | 0.001*** |
| Systolic blood pressure | 0.73835 | -0.67441 | 0.0158 | 0.487 |
| eGFR | 0.09266 | -0.99570 | 0.0053 | 0.780 |
| CRP_us | 0.81536 | -0.57896 | 0.0667 | 0.025* |
| HbA1c | 0.50241 | -0.86463 | 0.0753 | 0.027* |
| HDL- cholesterol | -0.99377 | 0.11144 | 0.0963 | 0.016* |
| LDL-cholesterol | -0.91281 | 0.40839 | 0.0231 | 0.306 |
| triglycerides | 0.88166 | 0.47189 | 0.1560 | 0.001*** |
| Total fat mass in % | 0.96126 | -0.27565 | 0.2090 | 0.001*** |
| Visceral fat rating | 0.79766 | -0.60310 | 0.0719 | 0.030* |
| Nr of antidiabetics | 0.99683 | 0.07956 | 0.0541 | 0.069 |
| Nr of antihypertensives | -0.65734 | -0.75359 | 0.0008 | 0.964 |
| Nr of antihyperlipidemics | -0.07182 | 0.99742 | 0.0135 | 0.514 |
| White blood cells count | 0.87696 | -0.48055 | 0.2531 | 0.001*** |
| Age | -0.75498 | -0.65575 | 0.0653 | 0.034* |
| Bacterial quantity | -0.99870 | 0.05102 | 0.1788 | 0.001*** |
| Waist-to-hip-ratio | -0.24996 | -0.96826 | 0.0860 | 0.015* |
| Diabetes Status _ No | -4.5303 | 5.4529 | 0.0630 | 0.002** |
| Diabetes Status _ Yes | 16.2677 | -19.5809 |  |  |
| Sex_female | 4.2702 | 3.6557 | 0.0318 | 0.039* |
| Sex_male | -12.3180 | -10.5452 |  |  |
| Metformin intake_No | -4.0974 | -0.8525 | 0.0380 | 0.022* |
| Metformin intake_Yes | 25.4624 | 5.2977 |  |  |
| PPI intake No | -16.5911 | 7.6883 | 0.0674 | 0.001*** |
| PPI intake Yes | 9.5917 | -4.4448 |  |  |
| Timepoint_baseline | 20.2299 | 9.4054 | 0.2184 | 0.001*** |
| Timepoint_one_year | -31.7231 | -7.1606 |  |  |
| Timepoint_three_months | 16.9608 | 0.2972 |  |  |
| Metabolic syndrome No | -10.5658 | 7.8723 | 0.0886 | 0.001*** |
| Metabolic Syndrome Yes | 15.4622 | -11.5205 |  |  |
| T2D Alleviation_ Yes | 14.2149 | -2.4049 | 0.0278 | 0.068 |
| T2D Alleviation_No | -5.4523 | 0.9224 |  |  |
| EBL_ good responder | -4.0761 | 7.9539 | 0.0389 | 0.082 |
| EBL_ medium_responder | 9.7610 | -2.1739 |  |  |
| EBL_ poor_responder | 1.8058 | -13.0851 |  |  |
| Surgical procedure_RYGB | -0.6923 | 21.583 | 0.0147 | 0.220 |
| Surgical procedure_Sleeve | 56.641 | -176.590 |  |  |

**Supplementary Table 5:**
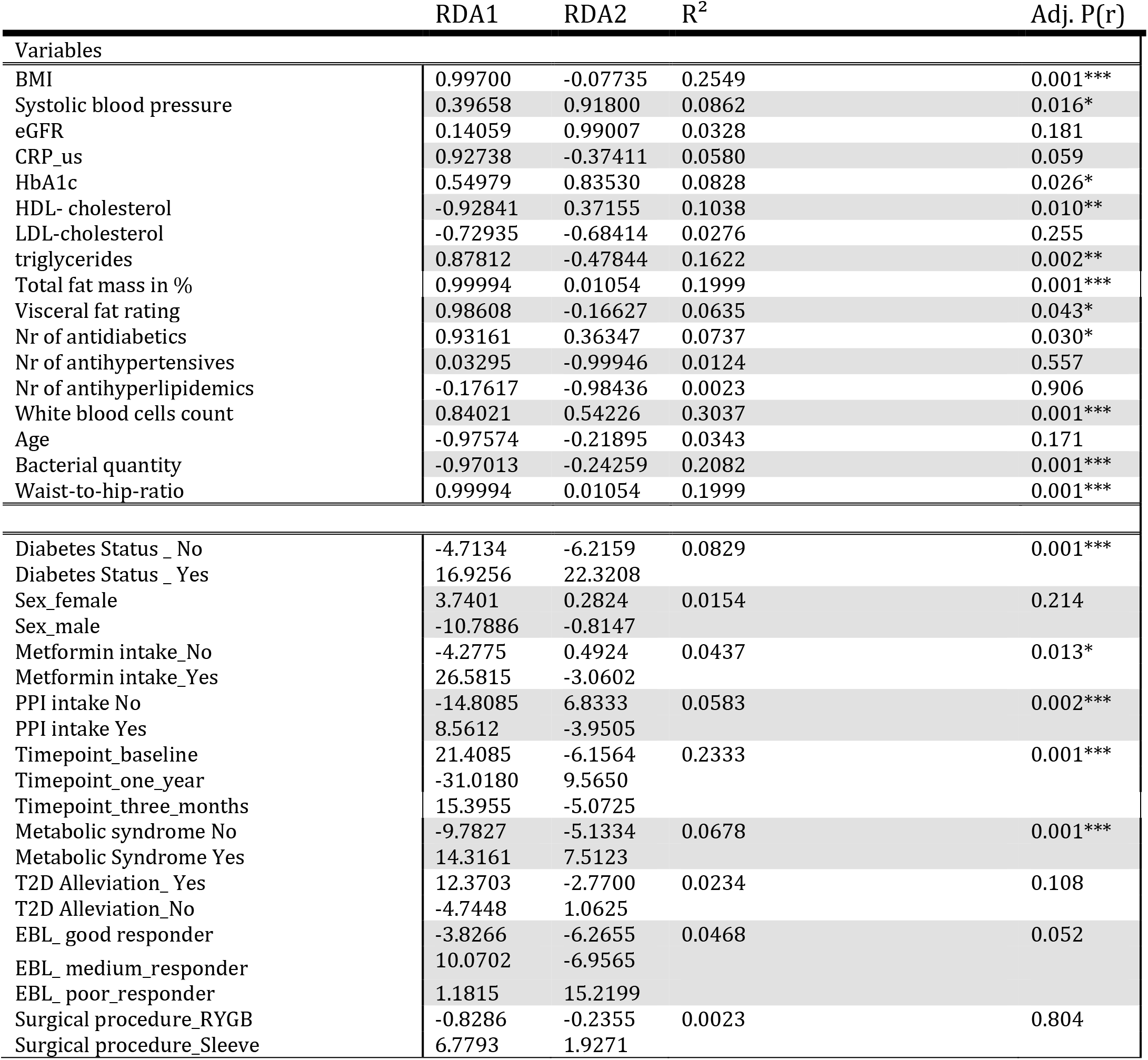
Envfit output for 27 variables on ASV level RDA

**Supplementary Table 6.** Spearman’s rank correlations between bacterial load and LBP with host variables in the replication cohort

| Covariate 1 | Covariate 2 | Beta-coefficient | P-value |
| --- | --- | --- | --- |
| Bacterial load in pg/μg isolated DNA | **Circ. TNF-α** | **-0.26** | **0.04** |
|  | LBP | -0.138 | 0.29 |
|  | HbA1c (%) | -0.12 | 0.33 |
|  | HOMA-IR | -0.20 | 0.11 |
|  | **Leukocytes count (in Gpt/l)** | **-0.38** | **<0.001** |
| LBP | **BMI** | **0.32** | **0.008** |
|  | **CRP** | **0.35** | **0.005** |
|  | **HOMA-IR** | **0.28** | **0.03** |
|  | HbA1c | 0.24 | 0.06 |
|  | **Leukocytes count** | **0.29** | **0.02** |

**Abbreviations:** **BMI:** Body mass index; **CRP:** C-reactive protein; **FPG:** Fasting plasma glucose; **HbA1c:** Glycated Hemoglobin A1c; **HOMA-IR:** Homeostatic model assessment for insulin resistance; **LBP:** Lipopolysaccharide binding protein, **TNF- $\alpha$ :** Tumor necrosis factor alpha

**Supplementary Table 7.**
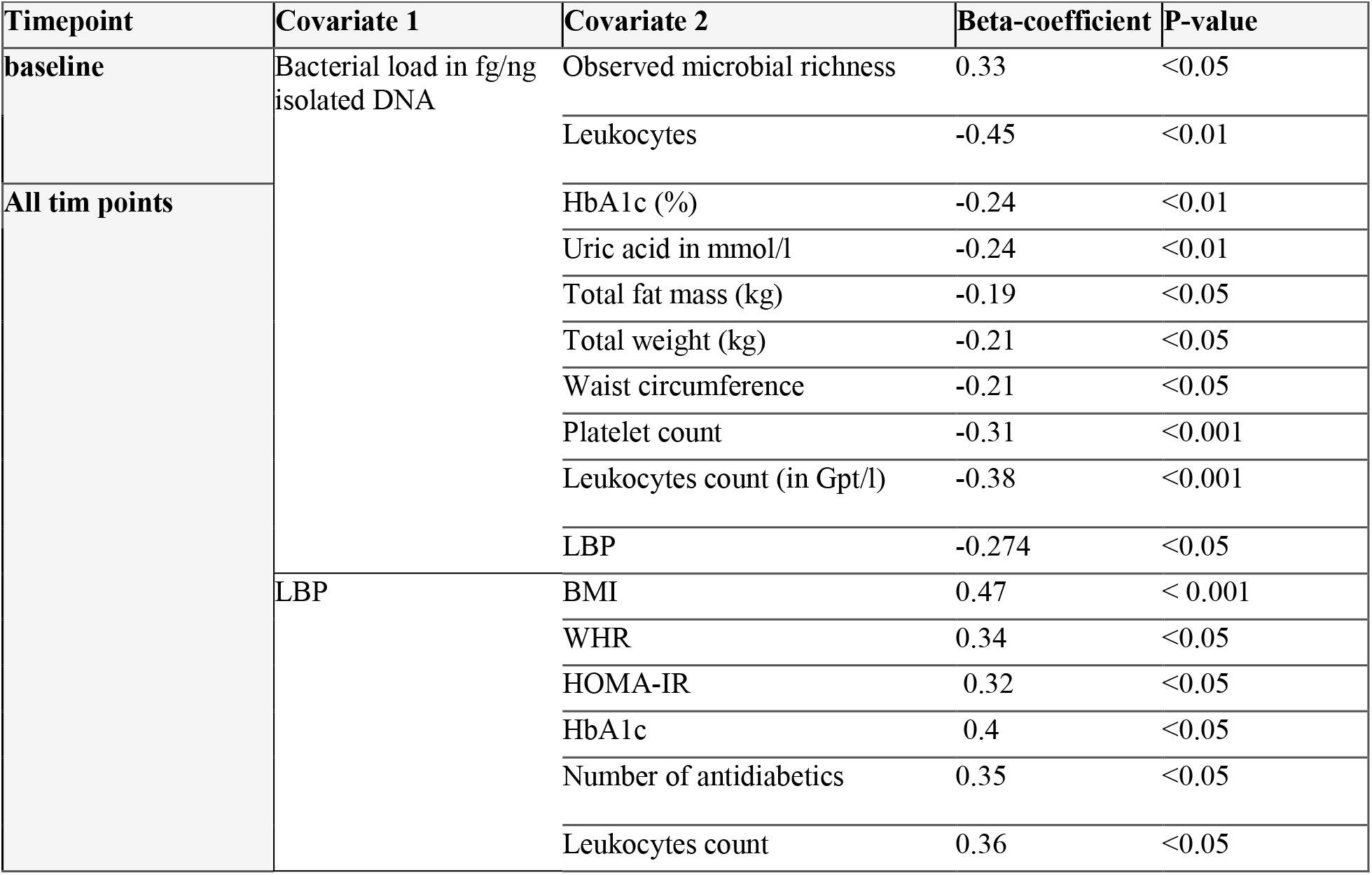
Spearman’s rank correlations between bacterial load and LBP with host variables in the study cohort

**Supplementary Table 8.**
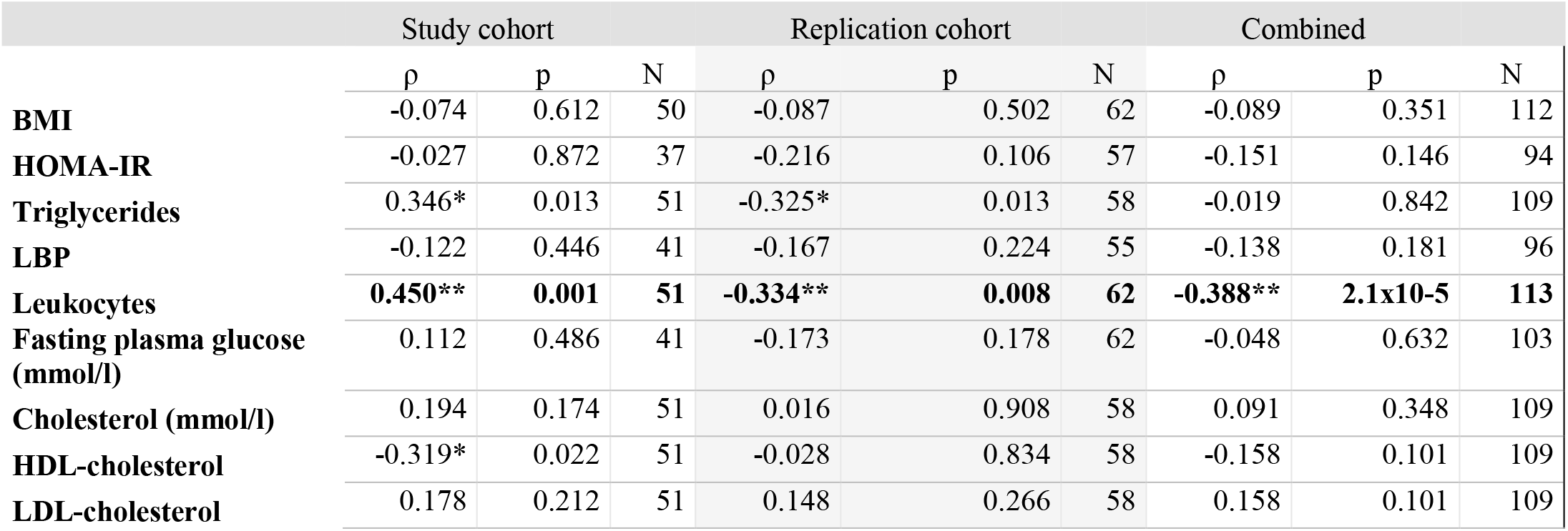
Spearman’s rank correlations between bacterial load and host variables in the study cohort at baseline and the replication cohort (for which only baseline is available)

|  | Study cohort |  |  | Replication cohort |  |  | Combined |  |  |
| --- | --- | --- | --- | --- | --- | --- | --- | --- | --- |
| | $\rho$ | p | N | $\rho$ | p | N | $\rho$ | p | N |
| **BMI** | -0.074 | 0.612 | 50 | -0.087 | 0.502 | 62 | -0.089 | 0.351 | 112 |
| **HOMA-IR** | -0.027 | 0.872 | 37 | -0.216 | 0.106 | 57 | -0.151 | 0.146 | 94 |
| **Triglycerides** | 0.346* | 0.013 | 51 | -0.325* | 0.013 | 58 | -0.019 | 0.842 | 109 |
| **LBP** | -0.122 | 0.446 | 41 | -0.167 | 0.224 | 55 | -0.138 | 0.181 | 96 |
| **Leukocytes** | **0.450\*\*** | **0.001** | **51** | **-0.334\*\*** | **0.008** | **62** | **-0.388\*\*** | **2.1x10-5** | **113** |
| **Fasting plasma glucose (mmol/l)** | 0.112 | 0.486 | 41 | -0.173 | 0.178 | 62 | -0.048 | 0.632 | 103 |
| **Cholesterol (mmol/l)** | 0.194 | 0.174 | 51 | 0.016 | 0.908 | 58 | 0.091 | 0.348 | 109 |
| **HDL-cholesterol** | -0.319* | 0.022 | 51 | -0.028 | 0.834 | 58 | -0.158 | 0.101 | 109 |
| **LDL-cholesterol** | 0.178 | 0.212 | 51 | 0.148 | 0.266 | 58 | 0.158 | 0.101 | 109 |

### Supplementary Figures Legends

**Supplementary Figure 1** Flowchart with number of subjects / Datasets and ASVs in each reported analyses

**Supplementary Figure 2** Number of ASVs with respective prevalence along Decontam scores: The contaminant scores show a bimodal distribution with low scores for lower prevalence ASVs around 0.1, higher prevalence ASVs (in which we are confident) start to rise beyond the threshold of 0.175.

**Supplementary Figure 3** flowchart showing T2D alleviation and response after bariatric surgery

**Supplementary Figure 4** correlation Triplot fitted site scores as linear combinations of the environmental variables and constraints (explanatory variables), highly significant variables with p< 0.002 are shown in red. First two canonical axes are visualized. Species scores are shown as red crosses, samples are shown as green triangles; Arrowheads depict significant explanatory variables and their directional contribution to the variance in the dataset.

**Supplementary Figure 5 (A)** Differentially abundant genera in subjects with T2D (n=9) compared to subjects without T2D (n=39) at 3 months post bariatric surgery. Differential abundance is calculated at ASV level using taxonomy after stringent control for contamination at a decontam score of 0.5 and reported at genus level. **(B)** Differentially abundant genera in subjects with T2D (n=4) compared to subjects without T2D (n=44) at one year post bariatric surgery. Differential abundance is calculated on ASV level using taxonomy after stringent control for contamination at a decontam score of 0.5 and reported at genus level. Color shading represents p-values. **(C)** Spearman’s rank correlations of relative selected genera abundance with host parameters at 3 months and one year post bariatric surgery. Selection included genera seen to be differentially abundant between groups (i.e. T2D, good/poor responders and T2D alleviation vs no T2D alleviation). Only genera are shown with at least one significant correlation with host markers. (+) refers to p-value < 0.05. Color represents correlation strength (Rho) according to color legend.

**Supplementary Figure 6 (A-D)** Spearman’s rank correlations of bacterial quantity in pg per total µg extracted DNA with markers or inflammation and metabolic disease over all timepoints.The grey area around regression line indicates the confidence interval at a confidence level of 95%.

**Supplementary Figure 7 (A-D)** Spearman’s rank correlations of bacterial quantity in pg per total µg extracted DNA with markers or inflammation and metabolic disease over all timepoints after removing statistical outliers for bacterial quantity. The grey area around regression line indicates the confidence interval at a confidence level of 95%.

**Supplementary Figure 8** bacterial quantity over time according to T2D and non T2D **(A,B)** respectively. Four samples with missing baseline bacterial quantity were eliminated. Boxplots are shown with Tukey- whiskers and mean (◆) as well as median. The three timepoints are compared using Kruskal-Wallis test and results are validated via Friedman’s test (Kruskal-Wallis p-value is depicted). Paired samples Wilcoxon signed-rank test is used to compare two groups at once.

**Supplementary Figure 9 (A)** Changes in LBP over time in good vs poor responders. **(B)** Changes in LBP over time on subjects with and without T2D at baseline **(C)** Changes in leukocytes over time in good vs poor responders. Boxplots are shown with Tukey-whiskers and mean (◆) as well as median. The three timepoints are compared using Kruskal-Wallis test and results are validated via Friedman’s test (Kruskal-Wallis p-value is depicted). Paired samples Wilcoxon signed-rank test is used to compare two groups at once.

**Supplementary Figure 10** Representative epifluorescence micrographs obtained by green light excitation and UV excitation of hybridized blood samples using CARD-FISH with HRP-labelled EUB338 probe (in red) and DAPI staining (in blue). (**A-B**) Positively hybridized bacterial cells (arrows) in fresh blood collected before mixed meal intake (**A**) and after mixed meal intake (**B**) from patient (5 years post bariatric surgery). (**C-D**) Healthy control blood samples collected before (C) and after (D) mixed meal intake shows no hybridized bacterial cells. Instead, the auto-fluorescence of blood cells can be observed upon increased exposure. (**E-F**) Control of CARD-FISH efficiency on blood samples collected from the healthy control before (**E**) and after (**F**) mixed meal intake and deliberately infected with Pseudomonas putida (Red) prior to hybridization shows positively hybridized cells of P. putida (arrows) as expected. In blue, DAPI staining of blood cells is observed. Scale bar 10 µm for all images

## Notes

### Competing Interest Statement

The authors have declared no competing interest.

### Author Declarations

Ethics approval and consent to participate This study was approved by the ethics committee of the University of Leipzig (application number: 047-13-28012013) and all participants involved gave written informed consent. The research was performed in accordance with the principles of the Declaration of Helsinki

