## Supplementary figures and images for "Far from the gut but still relevant? Circulating bacterial signature is linked to metabolic disease and shifts with metabolic alleviation after bariatric surgery"

### Supplementary Figure10

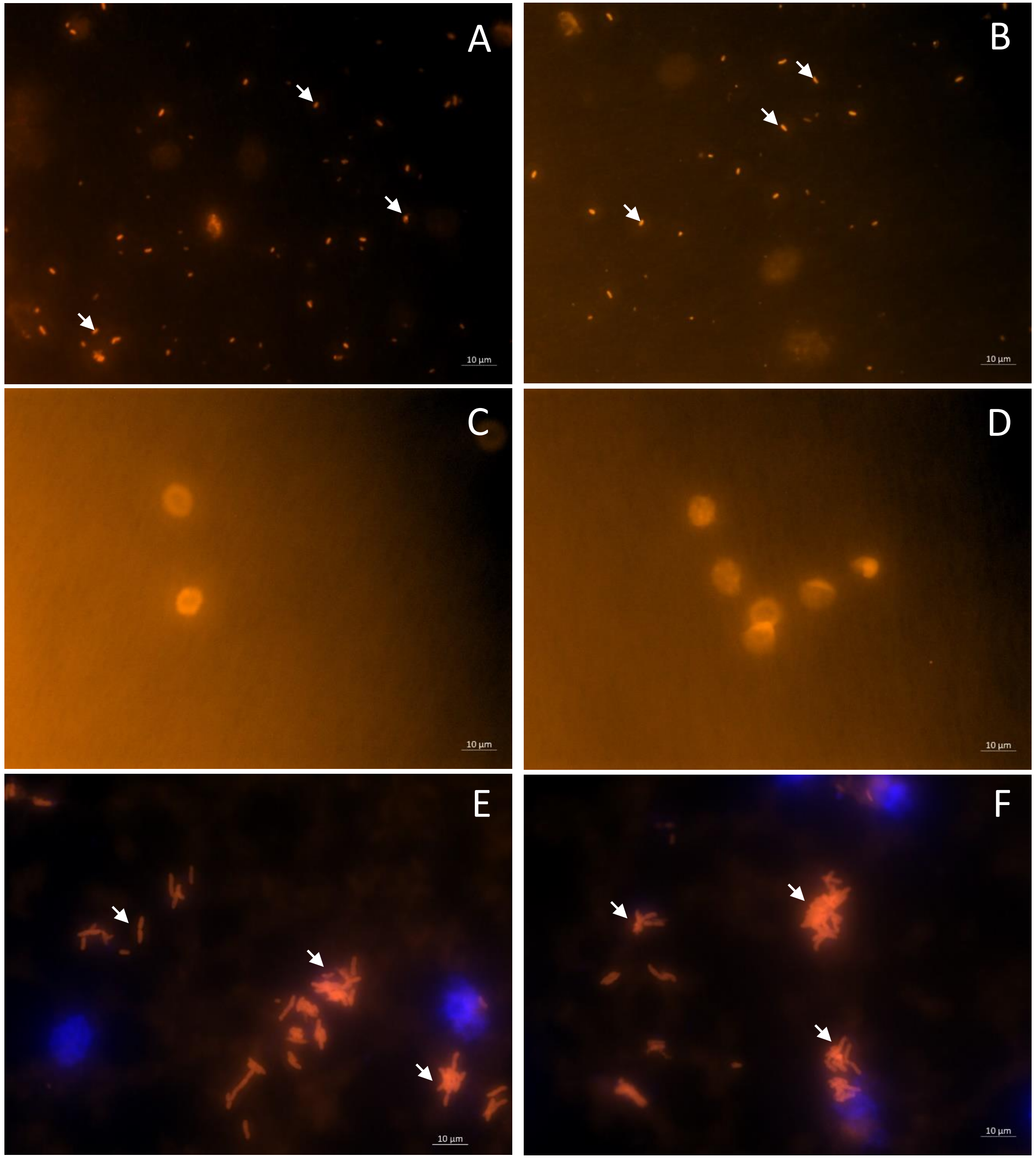

### Supplementary Figure 1

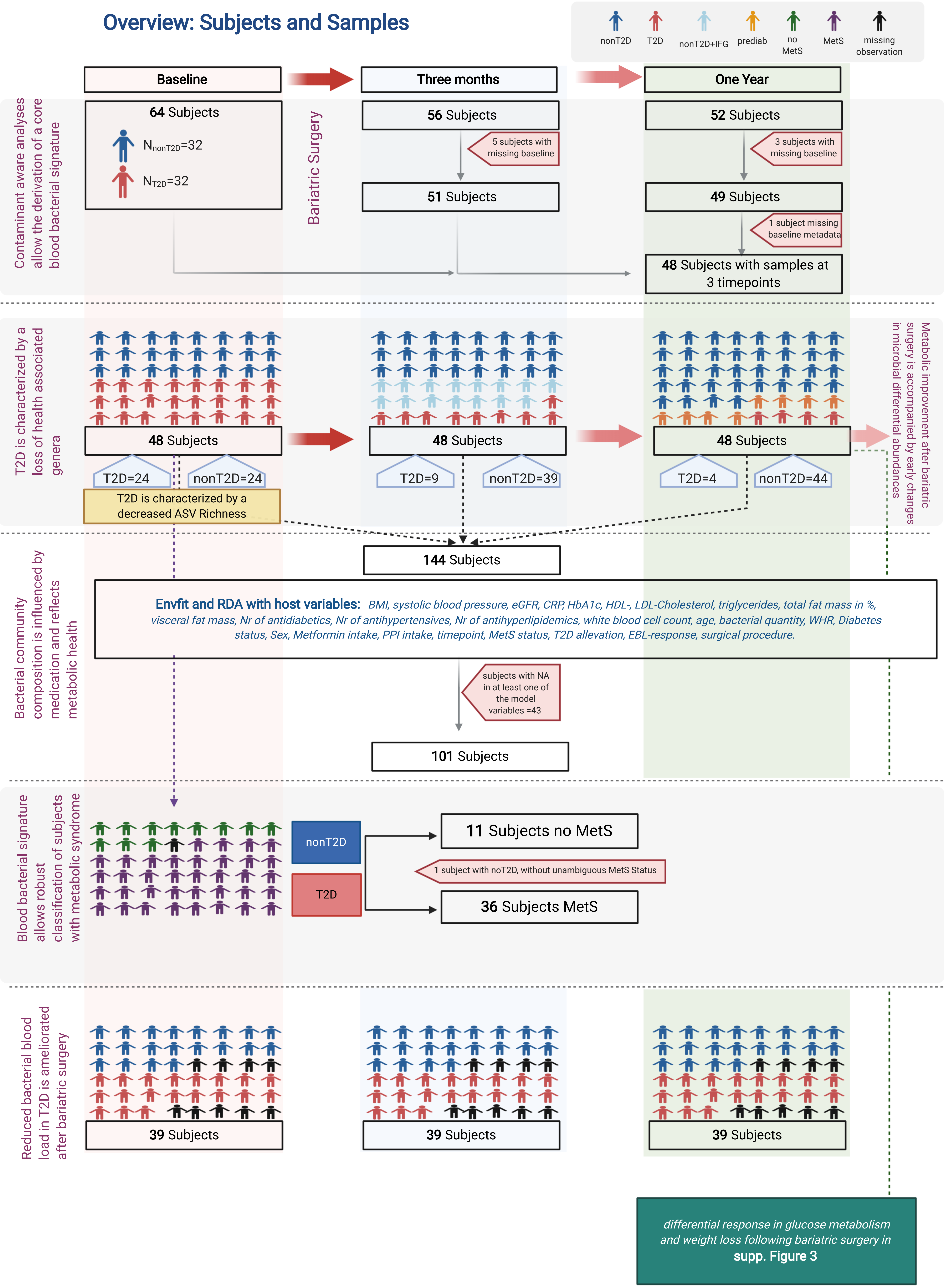

### Supplementary Figure 2

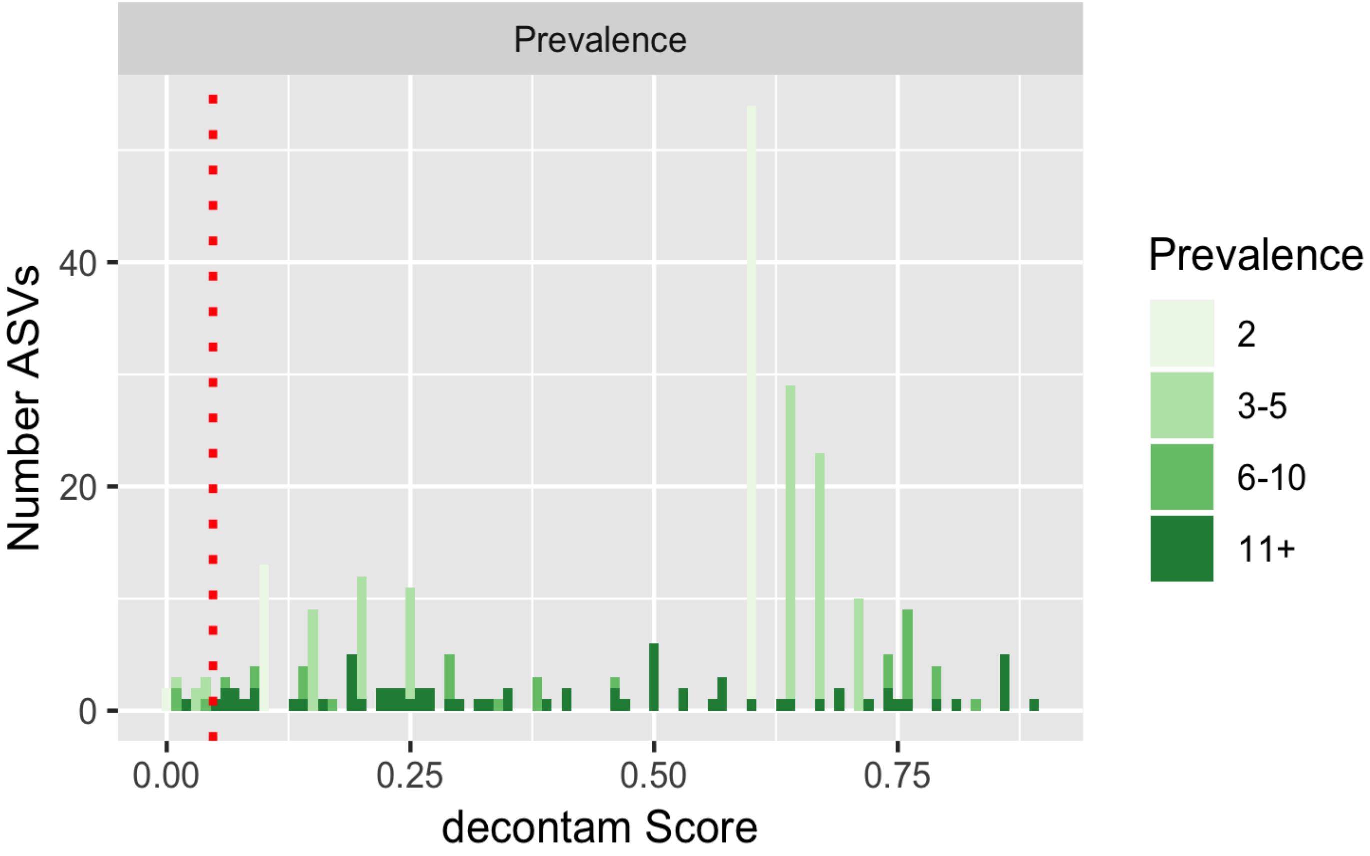

### Supplementary Figure 3

#### Differential Response in Glucose Metabolism and Weight loss Following Bariatric Surgery

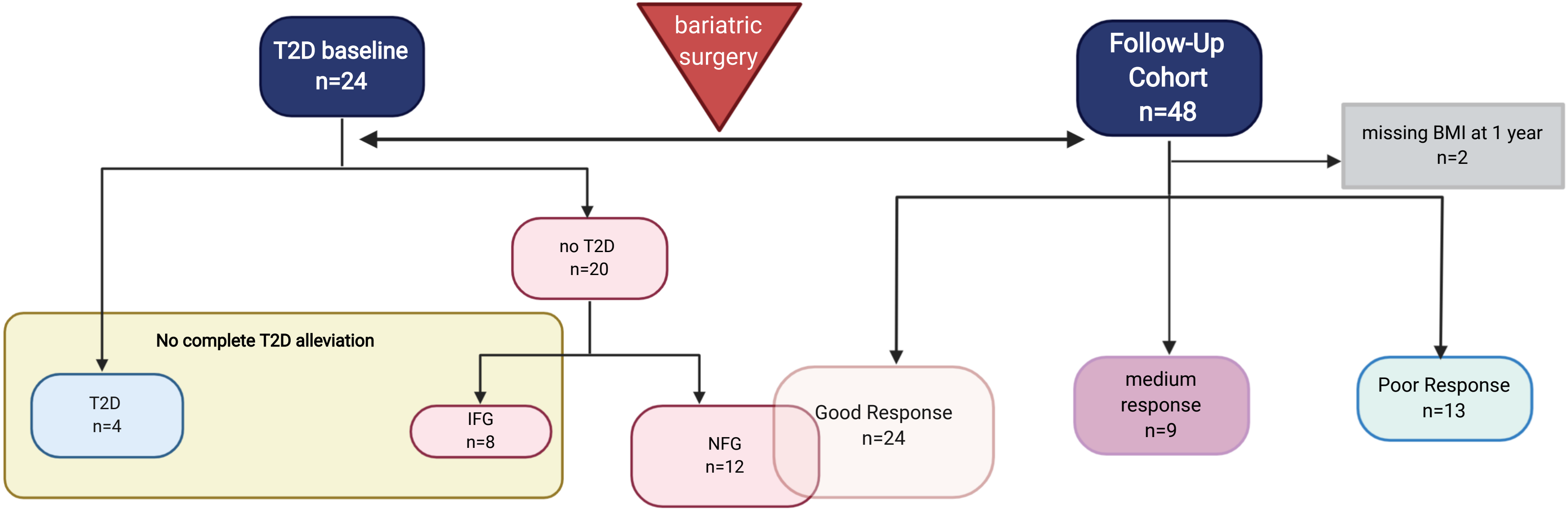

### Supplementary Figure 4

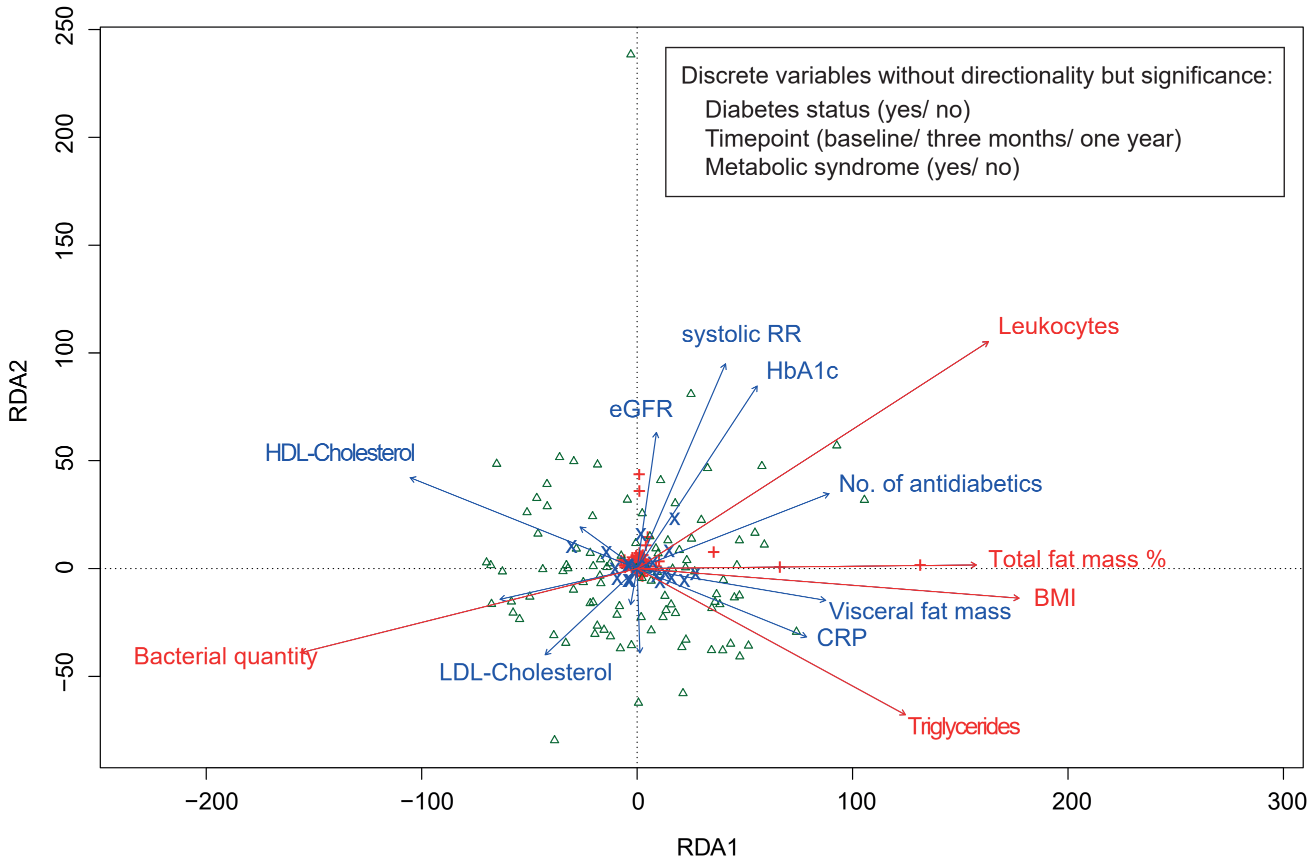

### Supplementary Figure 5

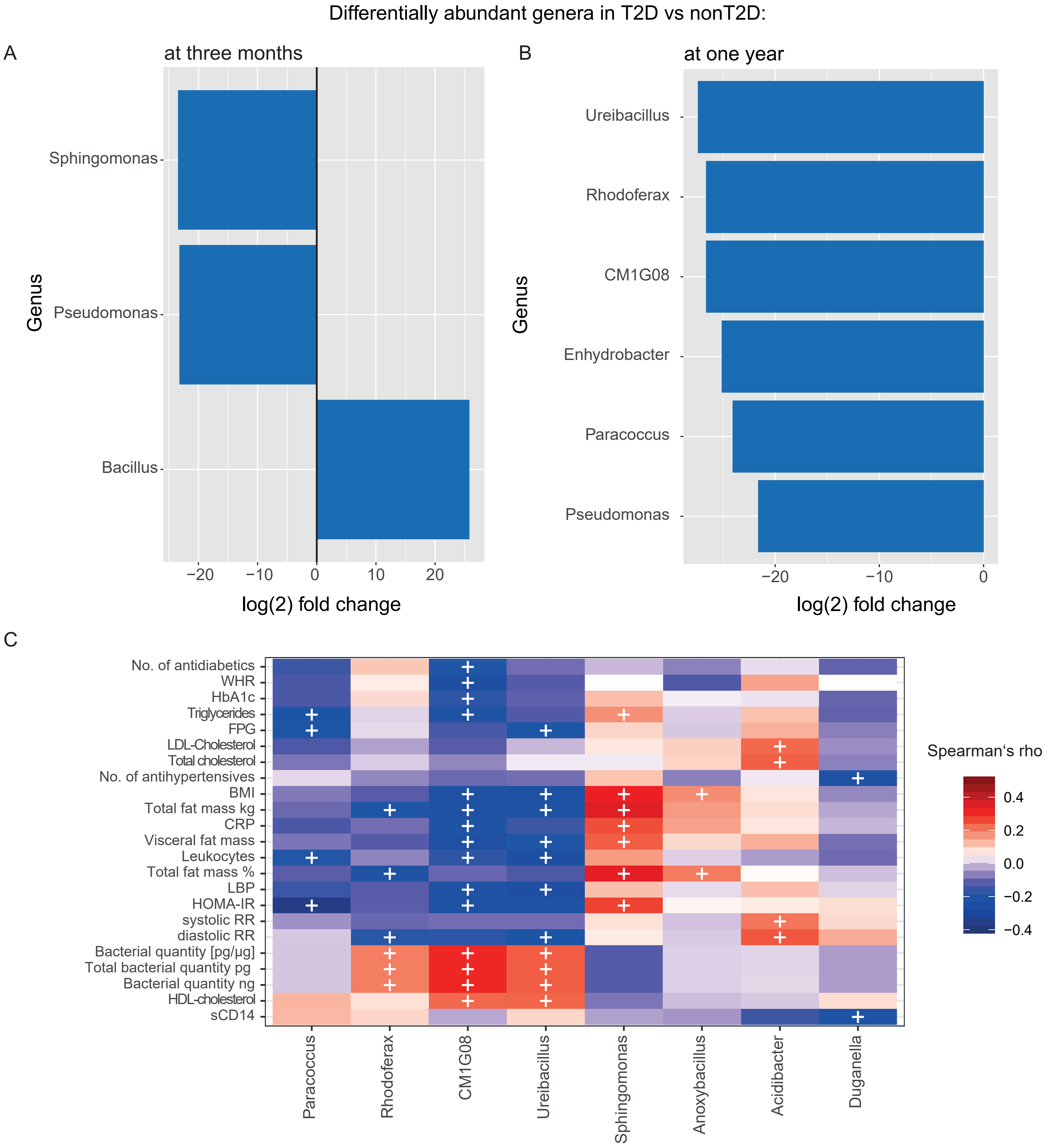

### Supplementary Figure 6

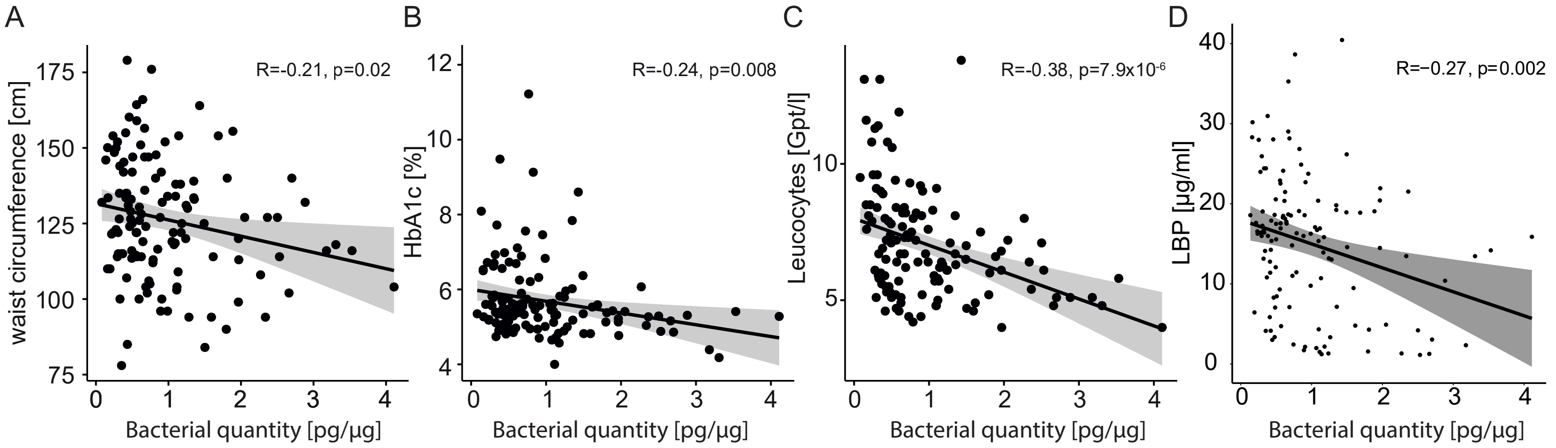

### Supplementary Figure 7

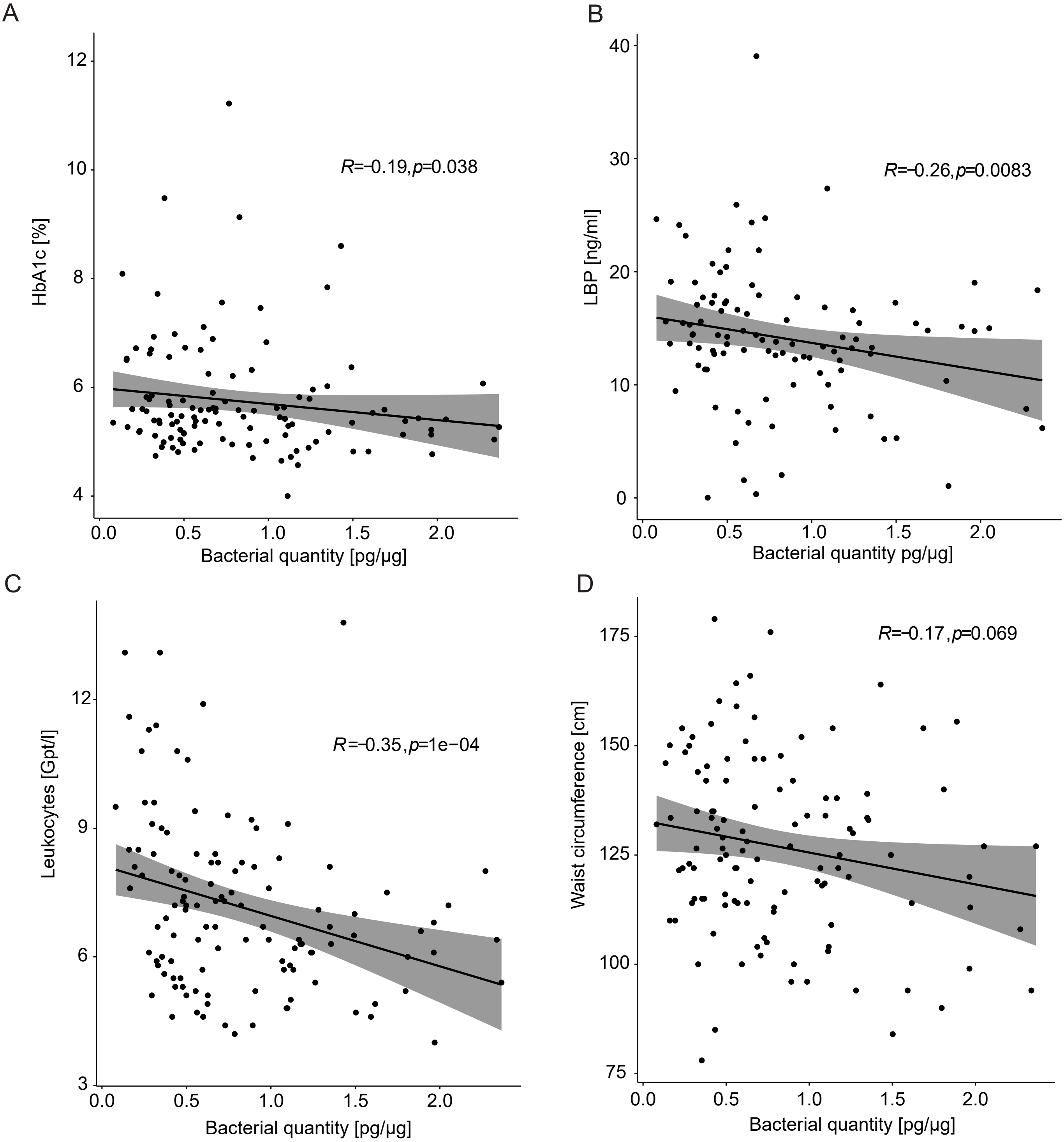

### Supplementary Figure 8

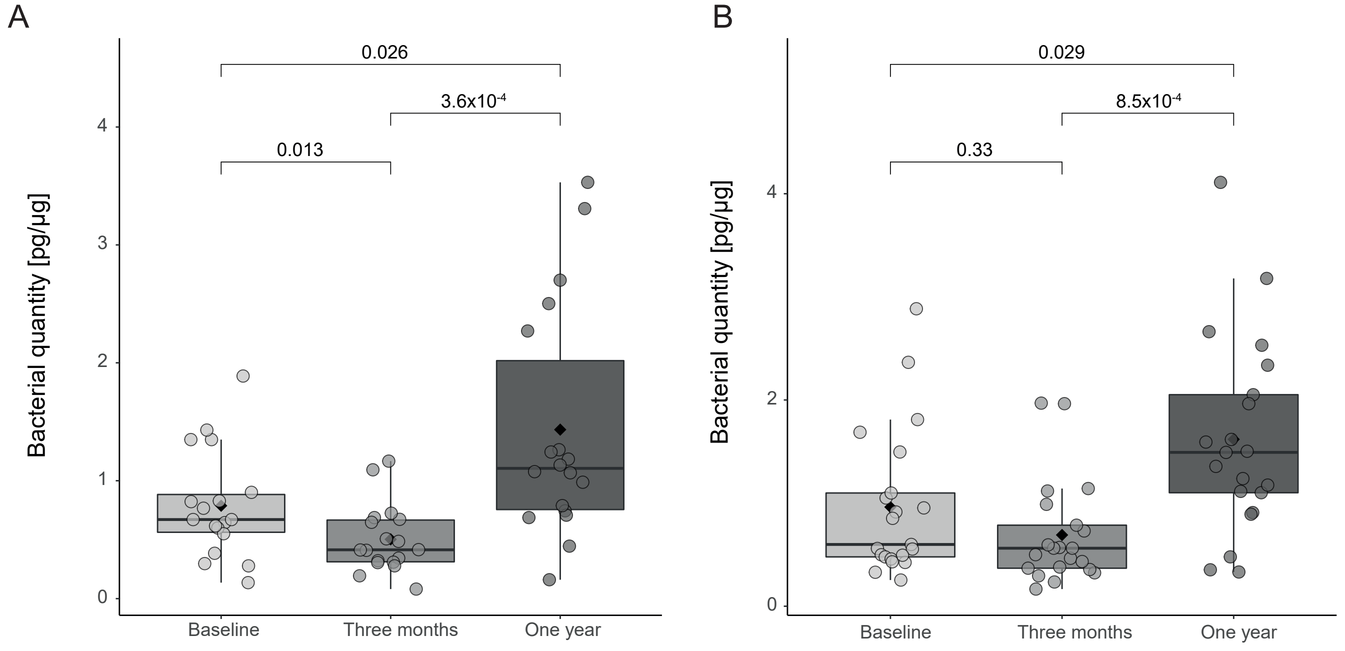

### Supplementary Figure 9

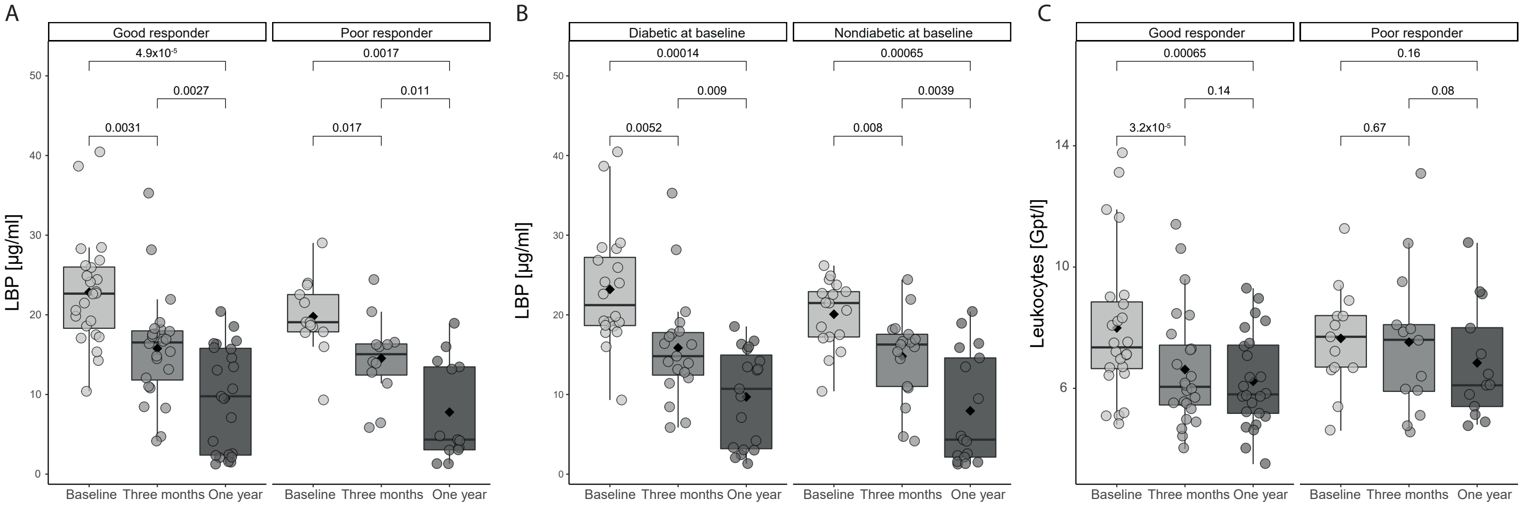
